# Prognostic Features in Surgically Resected Well-Differentiated Pancreatic Neuroendocrine Tumors: An Analysis of 904 Patients with 7882 Person-Years of Follow-Up

**DOI:** 10.1101/2025.04.01.25325055

**Authors:** Ashley L. Kiemen, Eric D. Young, Amanda L. Blackford, Pengfei Wu, Richard A. Burkhart, William R. Burns, John L. Cameron, Kelly Lafaro, Christopher Shubert, Zoe Gaillard, Uwakmfon-Abasi Ebong, Ian Reucroft, Yu Shen, Lucie Dequiedt, Valentina Matos, Günter Klöppel, Atsuko Kasajima, Jin He, Ralph H. Hruban

**Affiliations:** Department of Pathology, Sol Goldman Pancreatic Cancer Research Center, Johns Hopkins University School of Medicine, Baltimore, MD; Sidney Kimmel Comprehensive Cancer Center, Johns Hopkins University School of Medicine, Baltimore, MD; Department of Chemical and Biomolecular Engineering, Johns Hopkins University, Baltimore, MD; Institute for NanoBioTechnology, Johns Hopkins University.; Department of Functional Anatomy & Evolution, Johns Hopkins University School of Medicine, Baltimore, MD; Department of Oncology, Sol Goldman Pancreatic Cancer Research Center, Johns Hopkins University School of Medicine, Baltimore, MD; Department of Surgery, Sol Goldman Pancreatic Cancer Research Center, Johns Hopkins University School of Medicine, Baltimore, MD; Department of Pathology, TUM School of Medicine and Health, Technical University Munich, Munich, Germany

**Keywords:** Well-differentiated pancreatic neuroendocrine tumor, PanNET, Ki 67, grade, vascular invasion, stage, prognosis

## Abstract

**Importance:** The clinical behavior of well-differentiated pancreatic neuroendocrine tumors (PanNETs) is difficult to predict.

**Objective:** To define more accurately prognosticators for patients with a surgically resected PanNET.

**Design:** The pathology and Ki-67 immunolabeling index of PanNETs resected from 904 patients was correlated with patient outcome.

**Setting:** Academic tertiary care hospital.

**Participants:** Consecutive patients who had a PanNET resected between 1985 and 2025.

**Results:** The mean patient age at surgery was 56.6 years (SD 14.0), 477 were male (52.8%), and 7882 person-years of follow-up were obtained (mean 8.8 years, SD 6.5). The 10-year survival was 81% (95% CI: 77,86%) for patients with G1 PanNETs (Ki-67 <3%), 68% (95% CI: 61,76%) for patients with G2a PanNETs (Ki-67 3-<10%), 44% (95% CI: 29,66%) for patients with G2b PanNETs (Ki-67 of 10%-≤20%), and 23% (95% CI: 8,61%) for patients with G3 PanNETs. Metastases (HR 4.7, p <0.0001), vascular invasion (HR 3.0, p <0.0001), tumor size ≥ 2 cm (HR 2.88, p <0.0001), perineural invasion (HR 2.42, p<0.0001), and positive margins (HR 2.18, p <0.0001) were associated with worse overall survival. Insulinoma (HR 0.34, p=3e-04), sclerosing variant (HR 0.47, p=0.05), and cystic variant (HR 0.61, p=0.05) were associated with improved overall survival. T stage and N stage were all statistically significant classifiers of overall survival. Similar associations were found with respect to disease relapse. There was a significant (P<0.001) increase in the proportion of patients diagnosed with stage I vs stage IV disease over time.

**Conclusions and relevance:** This study supports the classification of PanNETs into four grades (G1, G2a, G2b, and G3) based on Ki-67 labeling, which allows a more accurate prognostic assessments of patients.

**Key Points:** *Question:* Does subdividing grade 2 well-differentiated pancreatic neuroendocrine tumors (PanNETs) into grade 2a (Ki-67 3-<10%), and grade 2b (Ki-67 10%-≤20%) improve patient prognostication after surgery?

*Findings:* In this single-institution cohort study of 904 adults, patients with grade 2a and patients with grade 2b PanNETs had distinct outcomes. Furthermore, the magnitude of the risk associated with metastases, perineural invasion, positive margins, tumor size, vascular invasion, sclerosing variant and cystic variant was refined.

*Meaning:* This study supports classifying PanNETs into four grades (G1, G2a, G2b, and G3), and provides a basis for accurate prognostic assessments of patients.

## Introduction

The incidence of well-differentiated pancreatic neuroendocrine tumors (PanNETs), the most prevalent non-ductal neoplasms of the pancreas, is increasing with increasing use of abdominal imaging.^1–7^ Clinical management of the growing numbers of patients with PanNETs is challenging.^8^ A number of new medical therapies are available for patients with advanced disease, and expert opinions vary on the optimal use of surgery, complicating the selection of the best therapy for patients.^8–10^ These challenges are perhaps greatest for patients with small, <2 cm, PanNETs, as prognostic classification of these relatively indolent tumors is imperfect, and the risks of surgery are not trivial.^11–14^ For example, Partelli and colleagues concluded, from the results of the ASPEN trial, that active surveillance is safe for patients with a small PanNET, while Lin and Huang, in an analysis of 1,102 patients, concluded that surgical resection is recommended for these patients.^12,14^ As a result of this uncertainty, guidelines vary, and the treatment of a significant proportion of patients is not based on any guidelines.^10,11^

A number of clinical, pathological and genetic prognostic markers for patients with a PanNET have been identified.^4,7^ Male sex and increasing age are both associated with shorter survival.^4,15^ Clinically, insulinomas and cystic PanNETs have been reported to be associated with improved survival.^7,16–20^ The pathological factors associated with survival include tumor size, lymph node status, metastasis, proliferation rate of the neoplastic cells, vascular invasion, and perineural invasion.^21–26^ Margin status, necrosis, a sclerotic growth pattern with serotonin expression, an invasive growth pattern, and the type and intensity of any associated immune infiltrates have also been reported as significant prognosticators.^21,27–34^ The genes *DAXX* and *ATRX* are frequently inactivated in PanNETs and inactivation of either one of these genes is associated with the alternative lengthening of telomeres (ALT) phenotype.^35^ The ALT phenotype and inactivation of *DAXX* or *ATRX* are also poor prognosticators.^36–39^ Despite these predictive features, prognostication remains imperfect, and there is still room for improvement.^7^

Recently, several studies have proposed refinements to prognostic features.^40,41^ Currently, PanNETs are divided into three grades based on proliferation rate of the neoplastic cells: G1 (Ki-67 <2%), G2 (Ki-67 3-20%) and G3 (Ki-67 >20%).^7,42^ Adsay and colleagues suggested that grade 2 PanNETs should be subdivided into two grades (G2a for Ki-67 3-<10%, and G2b for Ki-67 of 10%-≤20%), and that G2b PanNETs have a prognosis similar to G3 (Ki-67 >20%) PanNETs.^40^ For small tumors, <2 cm, Pawlik and colleagues suggested that vascular invasion may be a particularly useful prognosticator.^41^

To further refine the prognosticators for surgically resected PanNETs, particularly T1N0M0 tumors, and to specifically examine the most appropriate cut-off for proliferation rate, we reviewed the pathology of a large single-institution series of surgically resected PanNETs with up to three decades of follow-up and correlated findings with patient outcome. As it has been suggested that the incidence of clinically recognized PanNETs is increasing because of the incidental detection of asymptomatic lower-stage PanNETs, we also examined trends in age and stage at diagnosis over time.^1,43^

## Methods

### Study Population

This study was approved by the Institutional Review Board of the Johns Hopkins Hospital. The pathology and surgery files of the Johns Hopkins Hospital were searched for surgically resected PanNETs from January 1984 to January 2025. All available medical records were reviewed, as were all available microscope slides.

### Pathology Review

Five of the authors (ALK, EDY, GK, AK and RHH), as a group, reviewed all available slides. Neuroendocrine neoplasms metastatic to the pancreas, and neoplasms not meeting diagnostic definitions outlined in the 5th edition of the World Health Organization (WHO) Classification of Tumours of the Digestive System were excluded, l904 patients.^42^ Histologic slides were available from 883 of these 904 cases (Mean of 26, and median of 25 slides per case). Cystic PanNETs were defined by imaging and confirmed by gross appearance. Insulinoma was defined as a PanNET with associated clinical findings of hyperinsulinemic hypoglycemia.^7^ Patients had the classical “Whipple triad” of symptoms of hypoglycemia, low blood glucose levels (below 3.0 mmol per liter), and relief of symptoms when given glucose.^7,44^ Margins and size were recorded as reported in the pathology report. The sclerosing variant of PanNET was defined as a PanNET composed of cords of cells embedded in dense stromal fibrosis, centered on a large pancreatic duct, often with upstream ductal dilatation, and frequent expression of serotonin by immunolabeling.^32–34,45–47^

### Proliferation Rate

Tumor Ki-67 labeling index was available for 820 of the 904 patients. Ki-67 labeling was determined by one of two methods. For 302 of the cases, a labeling “hot spot” with the highest density of Ki-67 labeling cells was photographed, the photo printed and at least 500 neoplastic cells were manually counted.^48^ For 534 of the cases the percentage of neoplastic cells labeling in a “hot spot” was determined by the pathologist.^48^

### Statistics

The primary endpoint of this study was overall survival (OS), defined as the time from surgery to last follow-up or death and was estimated using the Kaplan-Meier method. Eleven of the 904 patients had more than one pancreas surgery. In these instances, we included their first surgery in the analyses. Twenty-nine of the 904 patients had <30 days of follow-up information and were excluded from recurrence and survival analyses. Hazard ratios (HR) for differences in OS according to patient subgroups were estimated with Cox proportional hazards models. Cause-specific survival was estimated similarly, except patients who died without relapse were censored on their date of death. Time to relapse was calculated as the time from surgery to date of relapse (event), death (competing event) or last follow-up, whichever came first. Patients with unknown disease status at last follow-up were excluded. The impact of margin status on outcome was calculated for the entire cohort and separately after excluding patients who had an enucleation procedure. Estimates of cumulative incidence of relapse at 2, 5, and 10 years and differences in time to relapse according to patient subgroups defined at the time of surgery were estimated using proportional sub-distribution hazards regression models.^49^

A multivariable model selection approach for OS was performed using a Least Absolute Shrinkage and Selection Operator (LASSO) method. LASSO, a regularization procedure, selects optimal covariates by applying a penalty factor through cross-validation. It performs both variable selection and shrinkage, reducing some coefficients to zero and improving model generalization by controlling overfitting.^50^ Age (per 5 years), gender, year of surgery, tumor size (per 1cm), Ki-67, TNM staging, and presence of insulinoma, sclerosing variant, cystic variant, vascular invasion, perineural invasion and margins were offered as candidates to the model selection. Estimates, 95% confidence intervals, and p-values were bias-corrected using the selective Inference package in R.^51^

Restricted cubic splines were used to assess the potential non-linear association between Ki-67 and tumor size with OS. To estimate changes in the prevalence of stage I disease over time, a multinomial regression model with stage (I, II, III, or IV) was fit with year of surgery included as the main independent variable, adjusting for age and gender. Due to the relatively small number of patients represented in some years, year of diagnosis was grouped into quintiles for this analysis. Predicted probabilities of being diagnosed with stage I disease were estimated from the model using the MNLpred package in R. Changes in age at surgery over time were estimated with a simple linear regression model. Analyses were completed with R version 4.4.1.^52^

## Results

### Patient Characteristics and Follow-Up

Patient characteristics are shown in Table 1. A total of 7882 person-years of follow-up were obtained (mean 8.8 years, SD 6.5) on the 875 patients followed for > 1 month. Of the 875 patients, 251 were followed until death. Disease status at last follow-up was known on 822 (94%) of the 875 patients.

**Table 1:** Descriptive summary of patient characteristics at the time of surgery and in follow-up.

| N = 904 |  | N = 904 |  |
| --- | --- | --- | --- |
| **Age at Surgery, years** |  | **Ki-67** |  |
| Mean (SD) | 56.6 (14.0) | Mean (SD) | 4.5 (6.6) |
| Median (Min, Max) | 58.0 [12.3, 93.3] | Median (Min, Max) | 2.0 [0.1, 58.6] |
| No. observations | 904 | No. observations | 820 |
| **Age at Surgery - no. (%)** |  | **Ki-67 - no. (%)** |  |
| < 60 | 509 (56.3) | < 2 | 289 (35.2) |
| 60+ | 395 (43.7) | 2+ | 531 (64.8) |
|  |  | No. Missing | 84 |
| **Gender - no. (%)** |  | **Grade - no. (%)** |  |
| Female | 427 (47.2) | G1 | 461 (56.2) |
| Male | 477 (52.8) | G2 | 335 (40.9) |
|  |  | G3 | 24 (2.9) |
| **Year of Surgery - no. (%)** |  | No. Missing | 84 |
| 1984 - 2000 | 106 (11.7) | **Grade, Proposed - no. (%)** |  |
| 2001 - 2010 | 306 (33.8) | G1 | 461 (56.2) |
| 2011 - 2020 | 390 (43.1) | G2a | 282 (34.4) |
| 2020 - 2024 | 102 (11.3) | G2b | 53 (6.5) |
|  |  | G3 | 24 (2.9) |
| **Years of Follow-Up** |  | No. Missing | 84 |
| Mean (SD) | 8.8 (6.5) | **Insulinoma - no. (%)** |  |
| Median (Min, Max) | 7.7 [0.0, 40.2] | No | 805 (89.0) |
| No. observations | 900 | Yes | 99 (11.0) |
| **Death or LFU < 30d of surgery - no. (%)** |  | **Functional - no. (%)** |  |
| Alive at 30d | 875 (97.2) | ACTH | 1 (0.8) |
| Death or LFU < 30d | 25 (2.8) | Gastrinoma | 15 (11.8) |
| No. Missing | 4 | Glucagonoma | 3 (2.4) |
| **Type of Surgery - no. (%)** |  | Insulinoma | 99 (78.0) |
| Whipple | 324 (35.8) | Pancreatic Polypeptide | 1 (0.8) |
| Total Pancreatectomy | 19 (2.1) | VIPoma | 8 (6.3) |
| Central Pancreatectomy | 13 (1.4) | No. Missing | 777 |
| Distal Pancreatectomy | 471 (52.1) | **Sclerosing Variant - no. (%)** |  |
| Enucleation | 76 (8.4) | No | 853 (94.4) |
| Hepatectomy | 0 (0.0) | Yes | 51 (5.6) |
| Biopsy Only | 0 (0.0) | **Cystic Variant - no. (%)** |  |
| No Surgery/Missing | 1 (0.1) | No | 813 (89.9) |
|  |  | Yes | 91 (10.1) |
| **Tumor Size, cm** |  | **Vascular Invasion - no. (%)** |  |
| Mean (SD) | 3.4 (3.0) | No | 667 (75.4) |
| Median (Min, Max) | 2.5 [0.1, 27.0] | Yes | 218 (24.6) |
| No. observations | 904 |  |  |
| **Tumor Size - no. (%)** |  |  |  |
| < 2cm | 338 (37.4) |  |  |
| 2cm + | 566 (62.6) |  |  |
| **T Stage - no. (%)** |  | No. Missing | 19 |
| T1 | 338 (37.4) | **Perineural Invasion - no. (%)** |  |
| T2 | 330 (36.5) | No | 601 (67.5) |
| T3 | 214 (23.7) | Yes | 290 (32.5) |
| T4 | 22 (2.4) | No. Missing | 13 |
| **N Stage - no. (%)** |  | **Margins - no. (%)** |  |
| N0 | 552 (61.1) | Negative | 814 (90.7) |
| Nx or Unknown | 83 (9.2) | Positive | 83 (9.3) |
| N1 | 269 (29.8) | No. Missing | 7 |
| **M Stage - no. (%)** |  | **Margins (Excluding Enucleations) - no. (%)** |  |
| M0 | 796 (88.1) | Negative | 768 (93.1) |
| M1a | 99 (11.0) | Positive | 57 (6.9) |
| M1b | 3 (0.3) | No. Missing | 79 |
| M1c | 6 (0.7) |  |  |
| **Stage Grouping - no. (%)** |  |  |  |
| I | 255 (28.2) |  |  |
| II | 263 (29.1) |  |  |
| III | 193 (21.3) |  |  |
| IV | 108 (11.9) |  |  |
| Unstaged | 85 (9.4) |  |  |

Cm=centimeter, d=days, max=maximum, min=minimum, no=number, SD=standard deviation

### Prognosticators in the Entire Cohort

OS for the entire cohort is shown in Figure 1A (sub-analysis of OS by stage in Supplemental Figure 1). Metastases (HR 4.71, 95% CI: 3.53, 6.28, p<0.0001), vascular invasion (HR 3.00, 95% CI: 2.31, 3.91, p<0.0001), tumor size ≥ 2 cm (HR 2.88, 95% CI: 2.10, 3.93, p<0.0001), perineural invasion (HR 2.42, 95% CI: 1.88, 3.12, p<0.0001), positive margins (HR 2.18, 95% CI: 1.56, 3.04, p<0.001 for the overall cohort and HR 3.1, 95% CI: 2.18,4.43, P≤0.0001 excluding enucleations), age ≥60 years (HR 2.08, 95% CI: 1.62, 2.68, p <0.0001), Ki-67 ≥ 2% (HR 1.74, 95% CI: 1.31, 2.30, p<0.0001) and male gender (HR 1.46, 95% CI: 1.14, 1.89, p=0.003) were all associated with worse OS (Table 2). Insulinoma (HR 0.34, 95% CI: 0.19, 0.62 p<0.001), sclerosing variant (HR 0.47, 95% CI: 0.22, 1.00, p=0.05), and cystic variant (HR 0.61, 95% CI: 0.38, 0.99 p=0.05) were associated with improved OS. Grade, T stage and N stage, as defined by the WHO, were also all statistically significant classifiers of outcome.^7,42^

**Figure 1:**
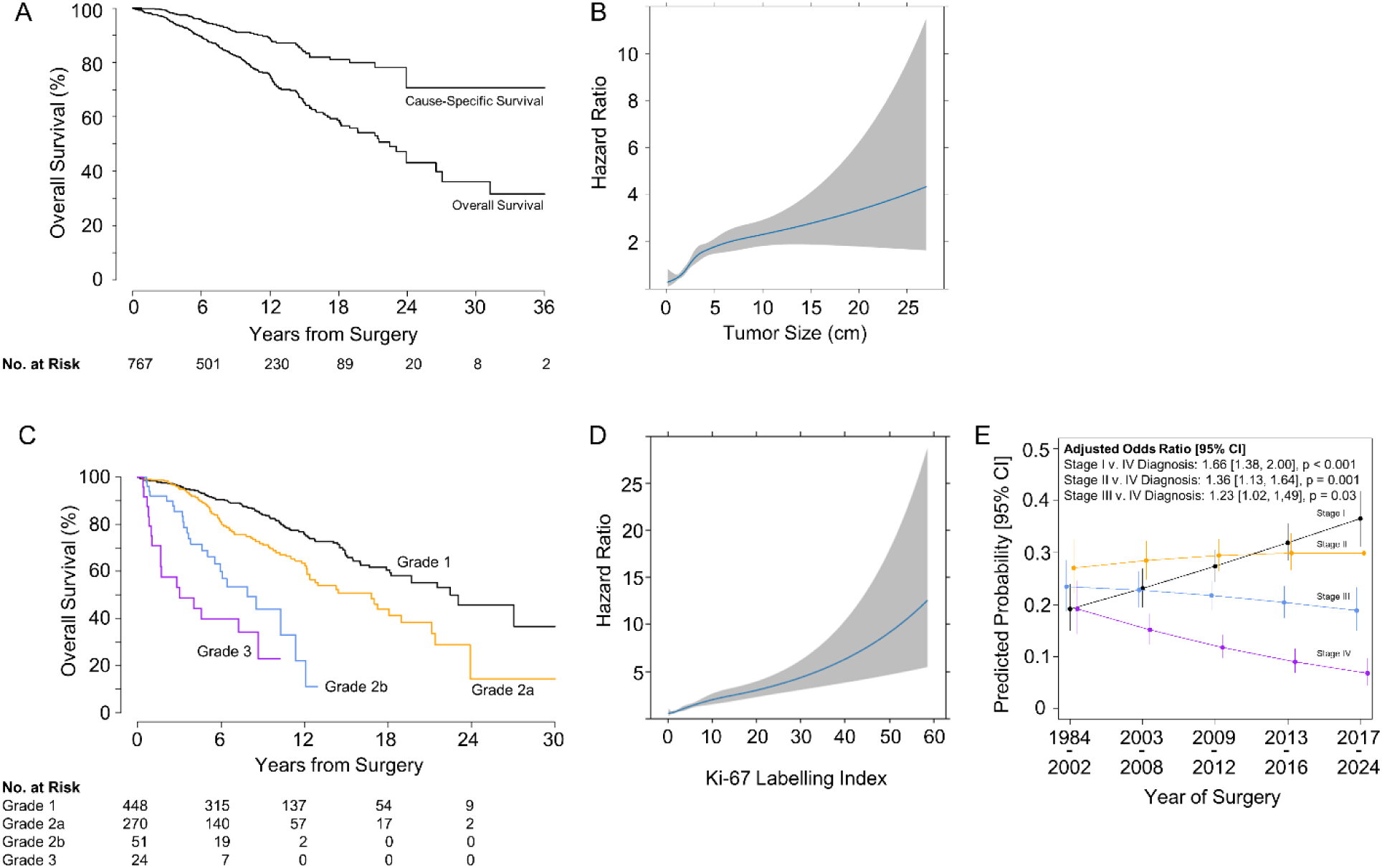
(a) Kaplan-Meier curve for overall survival for the entire cohort. (b) Changes in the hazard ratio for death with increases in tumor size, estimated with restricted cubic splines. The plotted results represent the hazard ratio for different values of the continuous predictor (tumor size), visualizing any non-linear effects captured by the restricted cubic spline. (c) Kaplan-Meier curves for overall survival, separately according to grade. (d) Changes in the hazard ratio for death with increases in Ki-67, estimated with restricted cubic splines. The plotted results represent the hazard ratio for different values of the continuous predictor (Ki-67), visualizing any non-linear effects captured by the restricted cubic spline. (e) Predicted probability [95% CI] of being diagnosed with stage I, II, III, or IV disease (y-axis) by year of surgery grouped into quintiles (x-axis). Probabilities and corresponding odds ratios for the average relative change in the odds of a stage I, II, or III diagnosis compared to stage IV with every increasing quintile of year of surgery, adjusted for age and sex, are estimated from a multinomial regression model.

**Table 2:**
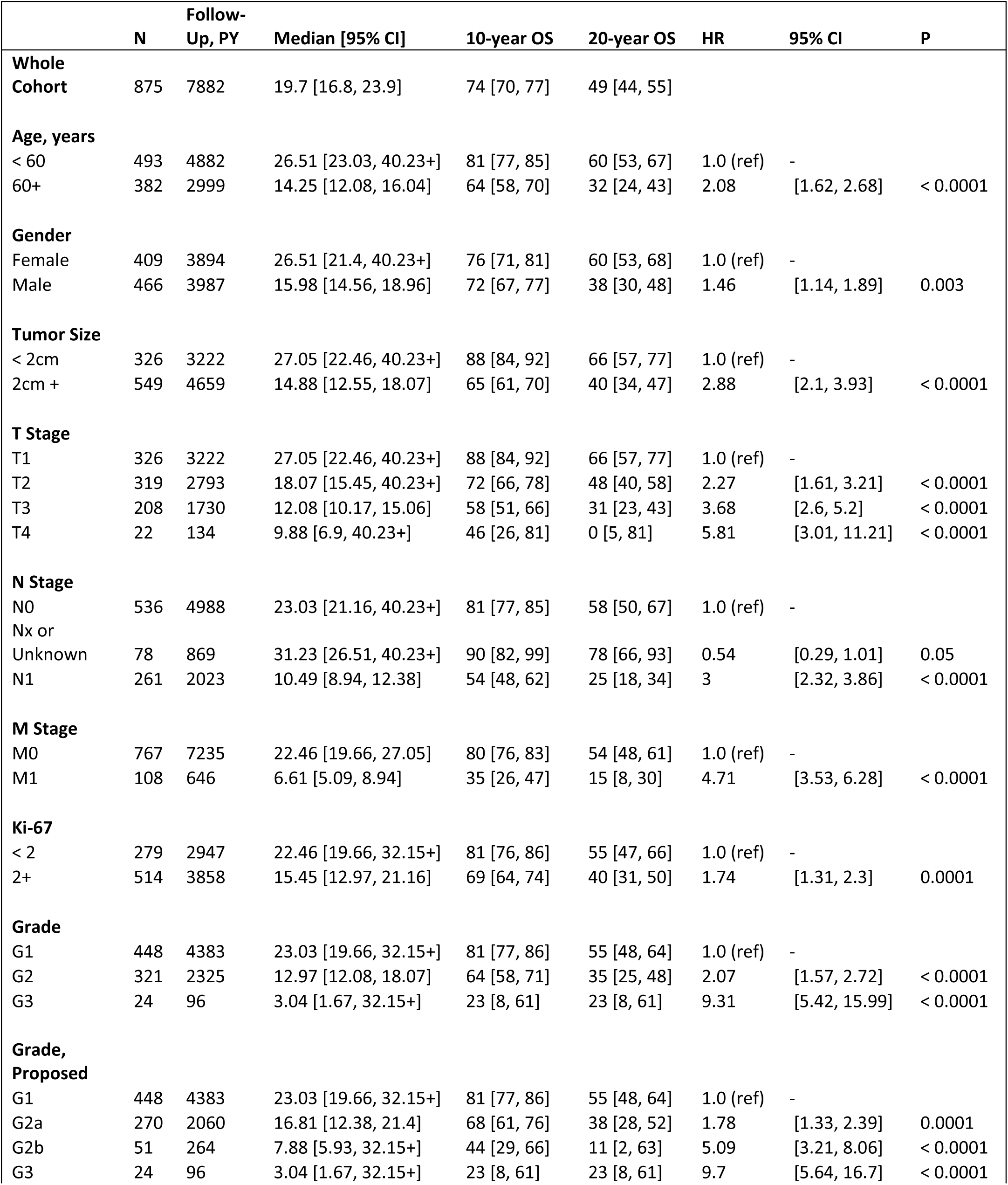
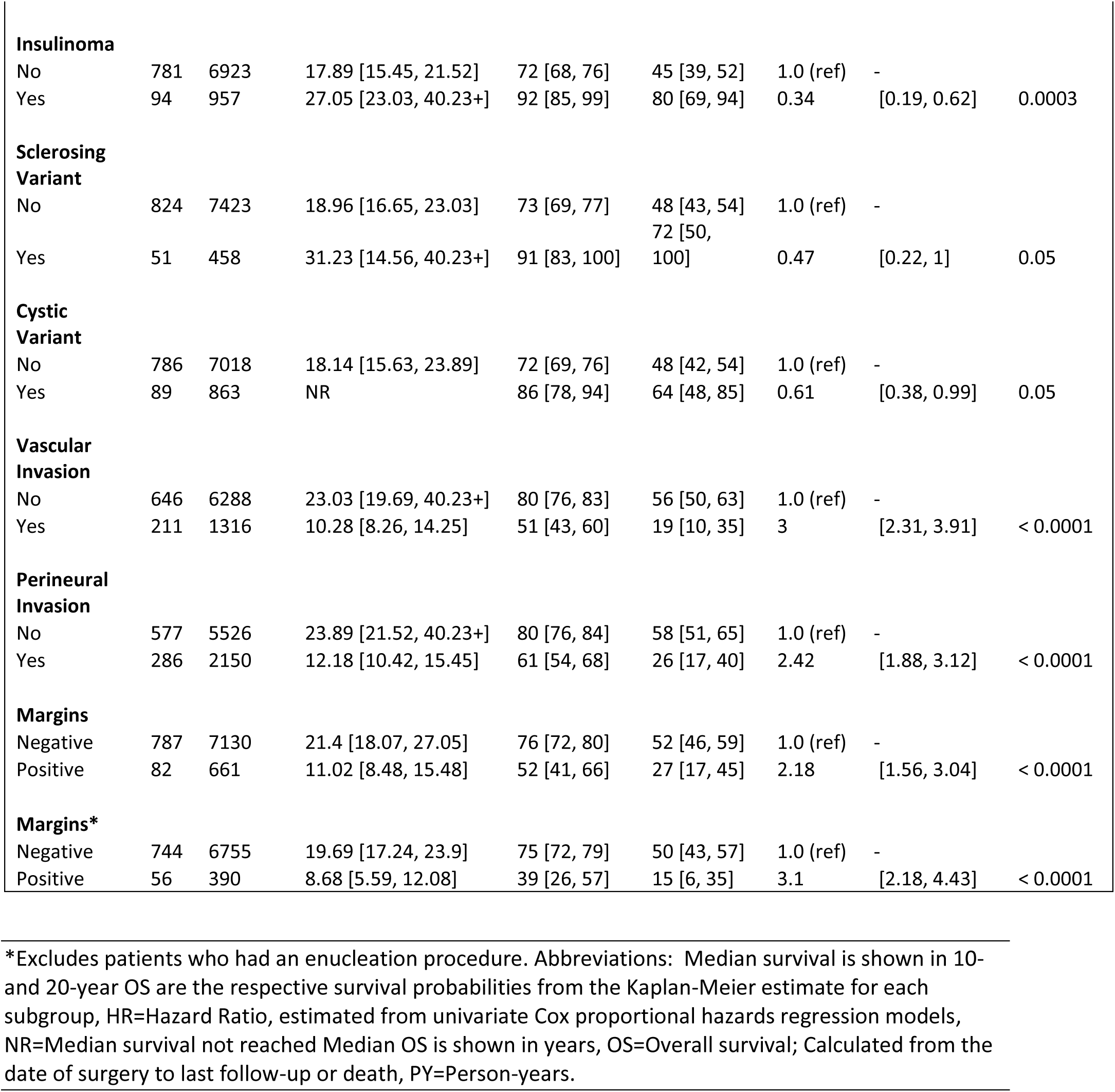
Overall Survival: Estimates of survival after surgery for the whole cohort and according to patient subgroups defined at the time of surgery.

|  | N | Follow-Up, PY | Median [95% CI] | 10-year OS | 20-year OS | HR | 95% CI | P |
| --- | --- | --- | --- | --- | --- | --- | --- | --- |
| **Whole Cohort** | 875 | 7882 | 19.7 [16.8, 23.9] | 74 [70, 77] | 49 [44, 55] |  |  |  |
| **Age, years** |  |  |  |  |  |  |  |  |
| < 60 | 493 | 4882 | 26.51 [23.03, 40.23+] | 81 [77, 85] | 60 [53, 67] | 1.0 (ref) | - |  |
| 60+ | 382 | 2999 | 14.25 [12.08, 16.04] | 64 [58, 70] | 32 [24, 43] | 2.08 | [1.62, 2.68] | < 0.0001 |
| **Gender** |  |  |  |  |  |  |  |  |
| Female | 409 | 3894 | 26.51 [21.4, 40.23+] | 76 [71, 81] | 60 [53, 68] | 1.0 (ref) | - |  |
| Male | 466 | 3987 | 15.98 [14.56, 18.96] | 72 [67, 77] | 38 [30, 48] | 1.46 | [1.14, 1.89] | 0.003 |
| **Tumor Size** |  |  |  |  |  |  |  |  |
| < 2cm | 326 | 3222 | 27.05 [22.46, 40.23+] | 88 [84, 92] | 66 [57, 77] | 1.0 (ref) | - |  |
| 2cm + | 549 | 4659 | 14.88 [12.55, 18.07] | 65 [61, 70] | 40 [34, 47] | 2.88 | [2.1, 3.93] | < 0.0001 |
| **T Stage** |  |  |  |  |  |  |  |  |
| T1 | 326 | 3222 | 27.05 [22.46, 40.23+] | 88 [84, 92] | 66 [57, 77] | 1.0 (ref) | - |  |
| T2 | 319 | 2793 | 18.07 [15.45, 40.23+] | 72 [66, 78] | 48 [40, 58] | 2.27 | [1.61, 3.21] | < 0.0001 |
| T3 | 208 | 1730 | 12.08 [10.17, 15.06] | 58 [51, 66] | 31 [23, 43] | 3.68 | [2.6, 5.2] | < 0.0001 |
| T4 | 22 | 134 | 9.88 [6.9, 40.23+] | 46 [26, 81] | 0 [5, 81] | 5.81 | [3.01, 11.21] | < 0.0001 |
| **N Stage** |  |  |  |  |  |  |  |  |
| N0 | 536 | 4988 | 23.03 [21.16, 40.23+] | 81 [77, 85] | 58 [50, 67] | 1.0 (ref) | - |  |
| Nx or Unknown | 78 | 869 | 31.23 [26.51, 40.23+] | 90 [82, 99] | 78 [66, 93] | 0.54 | [0.29, 1.01] | 0.05 |
| N1 | 261 | 2023 | 10.49 [8.94, 12.38] | 54 [48, 62] | 25 [18, 34] | 3 | [2.32, 3.86] | < 0.0001 |
| **M Stage** |  |  |  |  |  |  |  |  |
| M0 | 767 | 7235 | 22.46 [19.66, 27.05] | 80 [76, 83] | 54 [48, 61] | 1.0 (ref) | - |  |
| M1 | 108 | 646 | 6.61 [5.09, 8.94] | 35 [26, 47] | 15 [8, 30] | 4.71 | [3.53, 6.28] | < 0.0001 |
| **Ki-67** |  |  |  |  |  |  |  |  |
| < 2 | 279 | 2947 | 22.46 [19.66, 32.15+] | 81 [76, 86] | 55 [47, 66] | 1.0 (ref) | - |  |
| 2+ | 514 | 3858 | 15.45 [12.97, 21.16] | 69 [64, 74] | 40 [31, 50] | 1.74 | [1.31, 2.3] | 0.0001 |
| **Grade** |  |  |  |  |  |  |  |  |
| G1 | 448 | 4383 | 23.03 [19.66, 32.15+] | 81 [77, 86] | 55 [48, 64] | 1.0 (ref) | - |  |
| G2 | 321 | 2325 | 12.97 [12.08, 18.07] | 64 [58, 71] | 35 [25, 48] | 2.07 | [1.57, 2.72] | < 0.0001 |
| G3 | 24 | 96 | 3.04 [1.67, 32.15+] | 23 [8, 61] | 23 [8, 61] | 9.31 | [5.42, 15.99] | < 0.0001 |
| **Grade, Proposed** |  |  |  |  |  |  |  |  |
| G1 | 448 | 4383 | 23.03 [19.66, 32.15+] | 81 [77, 86] | 55 [48, 64] | 1.0 (ref) | - |  |
| G2a | 270 | 2060 | 16.81 [12.38, 21.4] | 68 [61, 76] | 38 [28, 52] | 1.78 | [1.33, 2.39] | 0.0001 |
| G2b | 51 | 264 | 7.88 [5.93, 32.15+] | 44 [29, 66] | 11 [2, 63] | 5.09 | [3.21, 8.06] | < 0.0001 |
| G3 | 24 | 96 | 3.04 [1.67, 32.15+] | 23 [8, 61] | 23 [8, 61] | 9.7 | [5.64, 16.7] | < 0.0001 |
| **Insulinoma** |  |  |  |  |  |  |  |  |
| No | 781 | 6923 | 17.89 [15.45, 21.52] | 72 [68, 76] | 45 [39, 52] | 1.0 (ref) | - |  |
| Yes | 94 | 957 | 27.05 [23.03, 40.23+] | 92 [85, 99] | 80 [69, 94] | 0.34 | [0.19, 0.62] | 0.0003 |
| **Sclerosing Variant** |  |  |  |  |  |  |  |  |
| No | 824 | 7423 | 18.96 [16.65, 23.03] | 73 [69, 77] | 48 [43, 54] | 1.0 (ref) | - |  |
| Yes | 51 | 458 | 31.23 [14.56, 40.23+] | 91 [83, 100] | 72 [50, 100] | 0.47 | [0.22, 1] | 0.05 |
| **Cystic Variant** |  |  |  |  |  |  |  |  |
| No | 786 | 7018 | 18.14 [15.63, 23.89] | 72 [69, 76] | 48 [42, 54] | 1.0 (ref) | - |  |
| Yes | 89 | 863 | NR | 86 [78, 94] | 64 [48, 85] | 0.61 | [0.38, 0.99] | 0.05 |
| **Vascular Invasion** |  |  |  |  |  |  |  |  |
| No | 646 | 6288 | 23.03 [19.69, 40.23+] | 80 [76, 83] | 56 [50, 63] | 1.0 (ref) | - |  |
| Yes | 211 | 1316 | 10.28 [8.26, 14.25] | 51 [43, 60] | 19 [10, 35] | 3 | [2.31, 3.91] | < 0.0001 |
| **Perineural Invasion** |  |  |  |  |  |  |  |  |
| No | 577 | 5526 | 23.89 [21.52, 40.23+] | 80 [76, 84] | 58 [51, 65] | 1.0 (ref) | - |  |
| Yes | 286 | 2150 | 12.18 [10.42, 15.45] | 61 [54, 68] | 26 [17, 40] | 2.42 | [1.88, 3.12] | < 0.0001 |
| **Margins** |  |  |  |  |  |  |  |  |
| Negative | 787 | 7130 | 21.4 [18.07, 27.05] | 76 [72, 80] | 52 [46, 59] | 1.0 (ref) | - |  |
| Positive | 82 | 661 | 11.02 [8.48, 15.48] | 52 [41, 66] | 27 [17, 45] | 2.18 | [1.56, 3.04] | < 0.0001 |
| **Margins\*** |  |  |  |  |  |  |  |  |
| Negative | 744 | 6755 | 19.69 [17.24, 23.9] | 75 [72, 79] | 50 [43, 57] | 1.0 (ref) | - |  |
| Positive | 56 | 390 | 8.68 [5.59, 12.08] | 39 [26, 57] | 15 [6, 35] | 3.1 | [2.18, 4.43] | < 0.0001 |

\*Excludes patients who had an enucleation procedure. Abbreviations: Median survival is shown in 10- and 20-year OS are the respective survival probabilities from the Kaplan-Meier estimate for each subgroup, HR=Hazard Ratio, estimated from univariate Cox proportional hazards regression models, NR=Median survival not reached Median OS is shown in years, OS=Overall survival; Calculated from the date of surgery to last follow-up or death, PY=Person-years.

Non-linear models for the association between tumor size and survival revealed a stronger positive association between 0 and 4 cm that reduces but remains positive without an additional inflection point for tumors >4 cm (Figure 1B).

### Subdividing Grade 2 PanNETs

We found that patients with G1 PanNETs had 10-year OS of 81% (95% CI: 77, 86%), G2a PanNETs 68% (95% CI: 61, 76%), G2b PanNETs 44% (95% CI: 29, 66%), and G3 PanNETs 23% (95% CI: 8, 61%) (Figure 1C and Table 2). Plotting Ki-67 versus the HR (Figure 1D) revealed a slight change to the slope of the curve at around a Ki-67 of 10%, supporting the introduction of a 10% threshold in the grading system.

### Prognosticators in M0, N0M0 and T1N0M0 Cohorts

As the most significant surgical decisions present in patients free of metastases at diagnosis, we estimated OS in lower stage subgroups, including patients who were M0 at surgery (Supplemental Table 1), N0M0 at surgery (Supplemental Table 2) and T1N0M0 at surgery (Table 3).

**Table 3:**
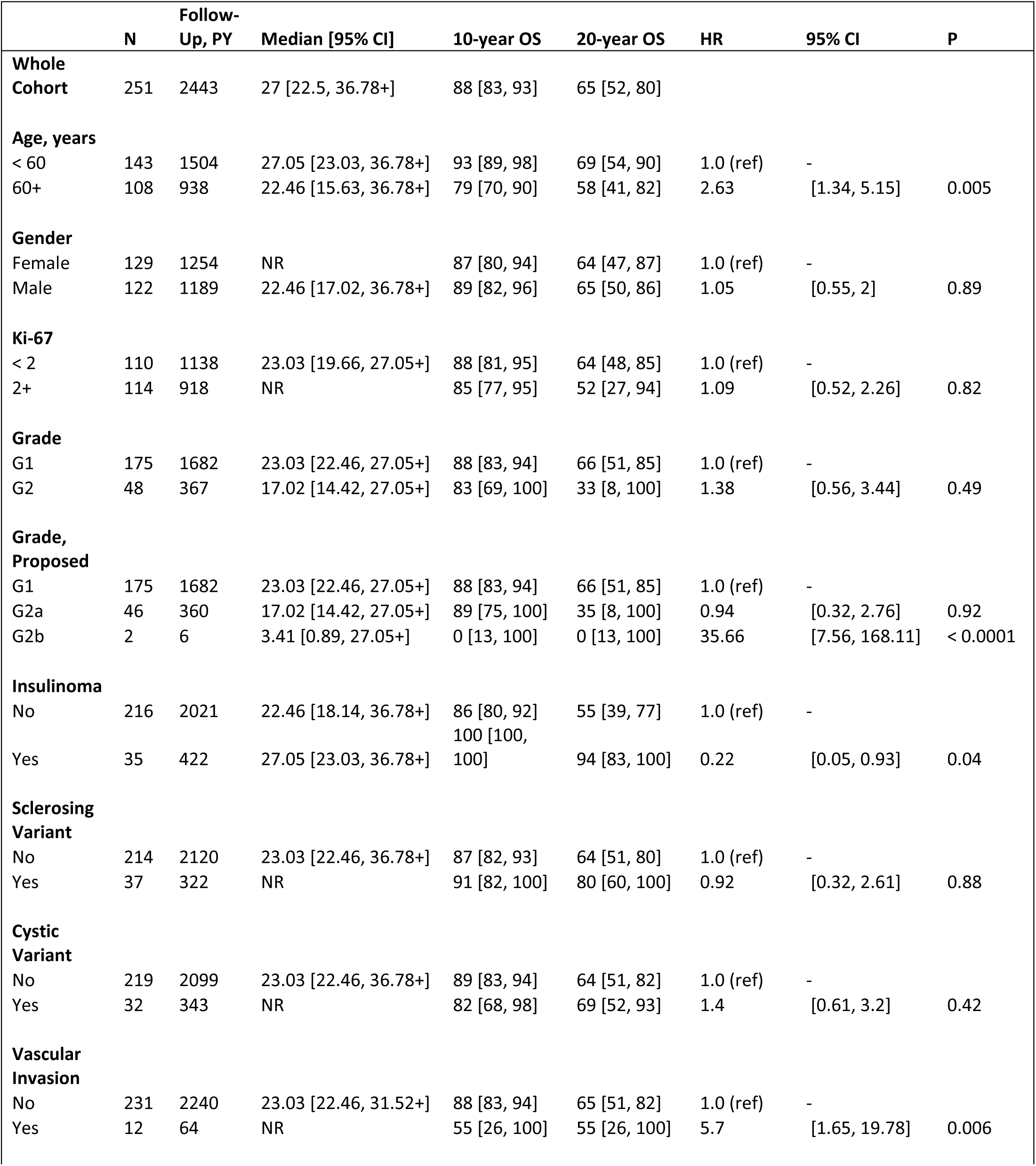
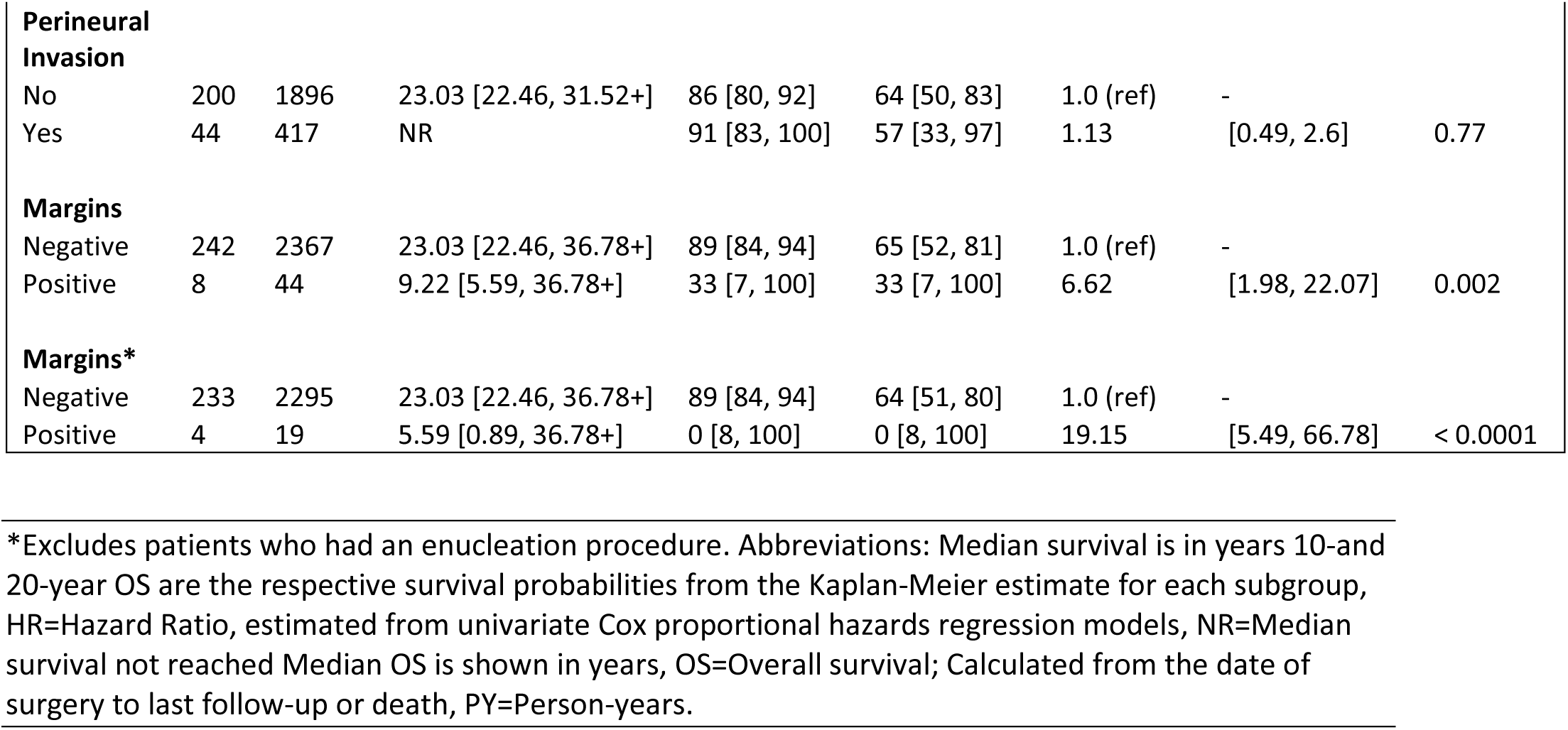
Overall Survival in Patients with T1N0M0 Disease: Estimates of survival after surgery for patients with M0, N0, and T1 disease, overall and according to patient subgroups defined at the time of surgery.

### Multivariate Analyses

Multivariate analyses of OS was performed on four cohorts (all patients, patients with M0 disease, patients with N0M0 disease, and patients with T1N0M0 disease) (Supplemental Table 3) using the LASSO method. For the whole cohort, M stage (M1 vs M0, HR 2.46, 95% CI: 1.85, 3.3, p<0.001), perineural invasion (HR 1.62, 95% CI: 1.24, 2.09, p=0.002), N stage (N1 vs N0, HR 1.56, 95% CI: 1.19, 2.02, p=0.005), age (per 5 years) (HR 1.26, 95% CI: 1.2, 1.32, p <0.001), Ki-67 (HR 1.06, 95% CI: 1.04, 1.09, p <0.001), tumor size (per 1cm) (HR 1.04, 95% CI: 1.01, 1.12, p=0.023), and year of surgery (per 1 year) (HR 0.96, 95% CI: 0.93, 0.97, p,0.001) were selected by the LASSO. Gender, vascular invasion and positive margins were also retained but not statistically significant. For the T1N0M0 cohort, age (HR 1.38, 95% CI: 1.18,1.59, p<0.001), year of surgery (HR 0.97, 95% CI: 0.91, 1.25, p = 0.556) and Ki-67 labeling (HR 1.15, 95% CI: 1.08,1.23, p<0.001) were retained.

### Cause-Specific Survival

Information on cause-specific survival for patients with M0 disease and M0N0 disease are presented in Supplemental Tables 4 and 5 respectively. With the exception of patient age (HR 1.29, 95% CI: 0.79, 2.11, p=0.31) and patient gender (HR 1.30, 95% CI: 0.80,2.10, p=0.30), all variables identified as statistically associated with OS remained significant. These results suggest that the patient age and gender findings observed with OS were due to the long follow-up obtained in this study, and not tumor-specific biological drivers.

### Cumulative Incidence of Relapse

Next, we estimated the cumulative probabilities of disease relapse at 2, 5, and 10 years after surgery for the M0, M0N0 and M0N0T1 cohorts (Table 4 and Supplemental Tables 6 and 7). In the M0 cohort, tumor size (HR 6.19, 95% CI: 3.9, 9.85, p<0.001), vascular invasion (HR 4.41, 95% CI: 3.18, 6.12, p<0.001), Ki-67 (HR 4.14, 95% CI: 2.65, 6.48, p<0.001), and perineural invasion (HR 3.17, 95% CI: 2.31, 4.35, p<0.001) remained poor prognosticators, while insulinoma (HR 0.26, 95% CI: 0.11, 0.63, p=0.003), sclerosing variant (HR 0.27, 95% CI: 0.09, 0.84, p=0.02), and cystic variant (HR 0.29, 95% CI: 0.13, 0.65, p=0.003) were associated with improved outcome. T stage and N stage were also significantly associated with relapse.

**Table 4:**
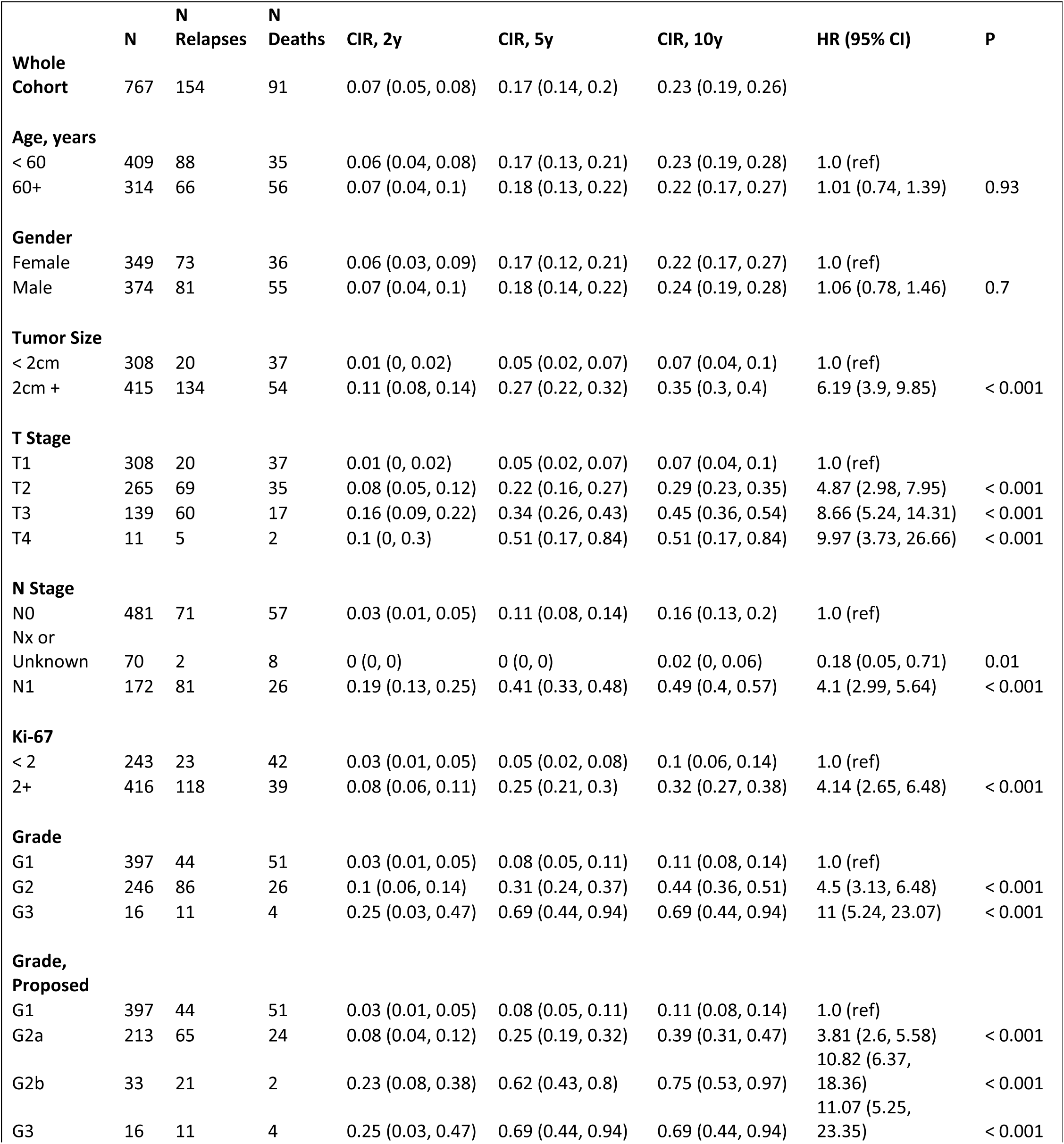
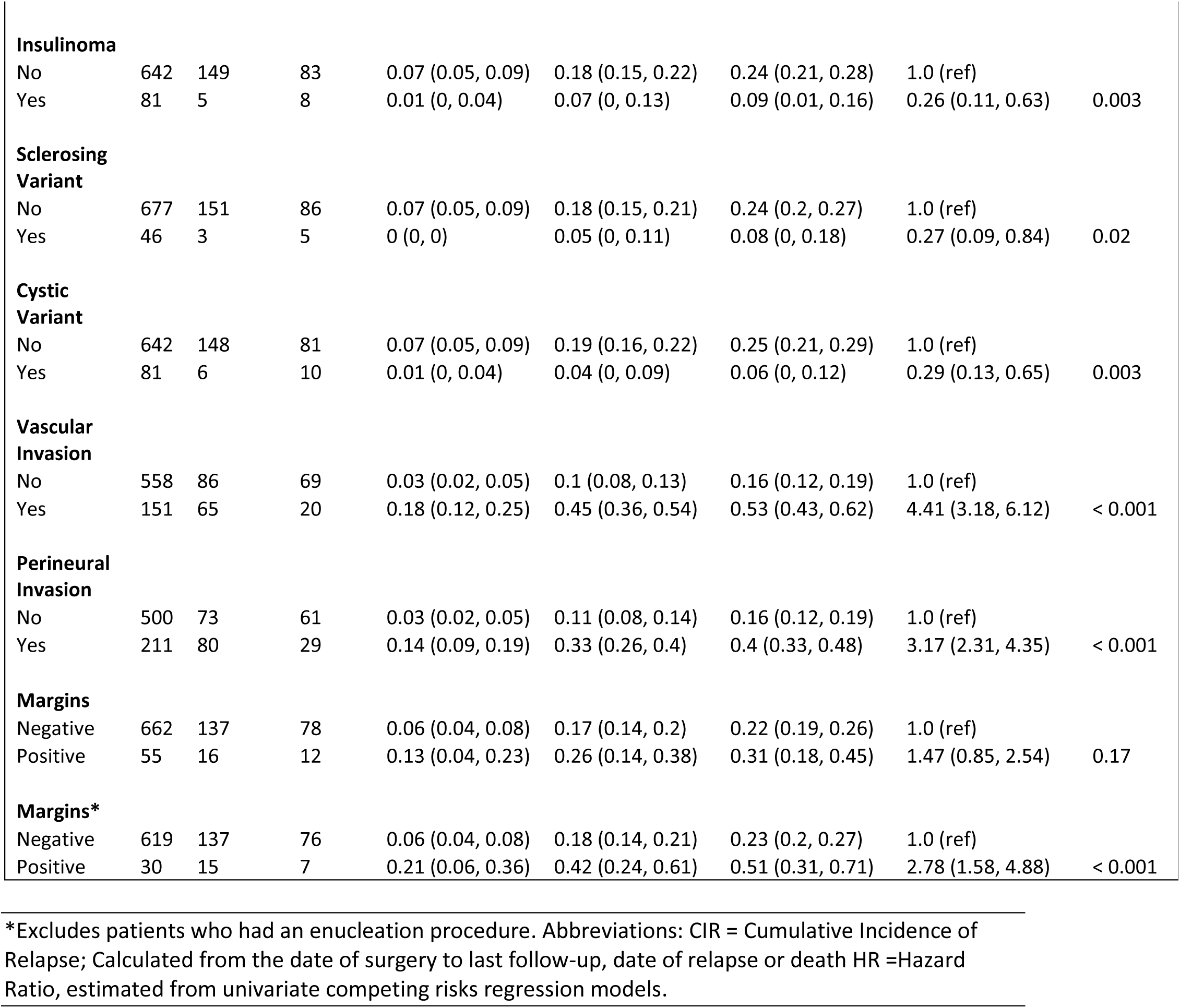
Cumulative Incidence of Relapse: Estimates of cumulative probability of relapse at 2, 5 and 10 years after surgery, accounting for death before relapse as a competing event, for the M0 cohort and according to patient subgroups defined at the time of surgery.

|  | N | N Relapses | N Deaths | CIR, 2y | CIR, 5y | CIR, 10y | HR (95% CI) | P |
| --- | --- | --- | --- | --- | --- | --- | --- | --- |
| **Whole Cohort** | 767 | 154 | 91 | 0.07 (0.05, 0.08) | 0.17 (0.14, 0.2) | 0.23 (0.19, 0.26) |  |  |
| **Age, years** |  |  |  |  |  |  |  |  |
| < 60 | 409 | 88 | 35 | 0.06 (0.04, 0.08) | 0.17 (0.13, 0.21) | 0.23 (0.19, 0.28) | 1.0 (ref) |  |
| 60+ | 314 | 66 | 56 | 0.07 (0.04, 0.1) | 0.18 (0.13, 0.22) | 0.22 (0.17, 0.27) | 1.01 (0.74, 1.39) | 0.93 |
| **Gender** |  |  |  |  |  |  |  |  |
| Female | 349 | 73 | 36 | 0.06 (0.03, 0.09) | 0.17 (0.12, 0.21) | 0.22 (0.17, 0.27) | 1.0 (ref) |  |
| Male | 374 | 81 | 55 | 0.07 (0.04, 0.1) | 0.18 (0.14, 0.22) | 0.24 (0.19, 0.28) | 1.06 (0.78, 1.46) | 0.7 |
| **Tumor Size** |  |  |  |  |  |  |  |  |
| < 2cm | 308 | 20 | 37 | 0.01 (0, 0.02) | 0.05 (0.02, 0.07) | 0.07 (0.04, 0.1) | 1.0 (ref) |  |
| 2cm + | 415 | 134 | 54 | 0.11 (0.08, 0.14) | 0.27 (0.22, 0.32) | 0.35 (0.3, 0.4) | 6.19 (3.9, 9.85) | < 0.001 |
| **T Stage** |  |  |  |  |  |  |  |  |
| T1 | 308 | 20 | 37 | 0.01 (0, 0.02) | 0.05 (0.02, 0.07) | 0.07 (0.04, 0.1) | 1.0 (ref) |  |
| T2 | 265 | 69 | 35 | 0.08 (0.05, 0.12) | 0.22 (0.16, 0.27) | 0.29 (0.23, 0.35) | 4.87 (2.98, 7.95) | < 0.001 |
| T3 | 139 | 60 | 17 | 0.16 (0.09, 0.22) | 0.34 (0.26, 0.43) | 0.45 (0.36, 0.54) | 8.66 (5.24, 14.31) | < 0.001 |
| T4 | 11 | 5 | 2 | 0.1 (0, 0.3) | 0.51 (0.17, 0.84) | 0.51 (0.17, 0.84) | 9.97 (3.73, 26.66) | < 0.001 |
| **N Stage** |  |  |  |  |  |  |  |  |
| N0 | 481 | 71 | 57 | 0.03 (0.01, 0.05) | 0.11 (0.08, 0.14) | 0.16 (0.13, 0.2) | 1.0 (ref) |  |
| Nx or Unknown | 70 | 2 | 8 | 0 (0, 0) | 0 (0, 0) | 0.02 (0, 0.06) | 0.18 (0.05, 0.71) | 0.01 |
| N1 | 172 | 81 | 26 | 0.19 (0.13, 0.25) | 0.41 (0.33, 0.48) | 0.49 (0.4, 0.57) | 4.1 (2.99, 5.64) | < 0.001 |
| **Ki-67** |  |  |  |  |  |  |  |  |
| < 2 | 243 | 23 | 42 | 0.03 (0.01, 0.05) | 0.05 (0.02, 0.08) | 0.1 (0.06, 0.14) | 1.0 (ref) |  |
| 2+ | 416 | 118 | 39 | 0.08 (0.06, 0.11) | 0.25 (0.21, 0.3) | 0.32 (0.27, 0.38) | 4.14 (2.65, 6.48) | < 0.001 |
| **Grade** |  |  |  |  |  |  |  |  |
| G1 | 397 | 44 | 51 | 0.03 (0.01, 0.05) | 0.08 (0.05, 0.11) | 0.11 (0.08, 0.14) | 1.0 (ref) |  |
| G2 | 246 | 86 | 26 | 0.1 (0.06, 0.14) | 0.31 (0.24, 0.37) | 0.44 (0.36, 0.51) | 4.5 (3.13, 6.48) | < 0.001 |
| G3 | 16 | 11 | 4 | 0.25 (0.03, 0.47) | 0.69 (0.44, 0.94) | 0.69 (0.44, 0.94) | 11 (5.24, 23.07) | < 0.001 |
| **Grade, Proposed** |  |  |  |  |  |  |  |  |
| G1 | 397 | 44 | 51 | 0.03 (0.01, 0.05) | 0.08 (0.05, 0.11) | 0.11 (0.08, 0.14) | 1.0 (ref) |  |
| G2a | 213 | 65 | 24 | 0.08 (0.04, 0.12) | 0.25 (0.19, 0.32) | 0.39 (0.31, 0.47) | 3.81 (2.6, 5.58) | < 0.001 |
| G2b | 33 | 21 | 2 | 0.23 (0.08, 0.38) | 0.62 (0.43, 0.8) | 0.75 (0.53, 0.97) | 10.82 (6.37, 18.36) | < 0.001 |
| G3 | 16 | 11 | 4 | 0.25 (0.03, 0.47) | 0.69 (0.44, 0.94) | 0.69 (0.44, 0.94) | 11.07 (5.25, 23.35) | < 0.001 |
| **Insulinoma** |  |  |  |  |  |  |  |  |
| No | 642 | 149 | 83 | 0.07 (0.05, 0.09) | 0.18 (0.15, 0.22) | 0.24 (0.21, 0.28) | 1.0 (ref) |  |
| Yes | 81 | 5 | 8 | 0.01 (0, 0.04) | 0.07 (0, 0.13) | 0.09 (0.01, 0.16) | 0.26 (0.11, 0.63) | 0.003 |
| **Sclerosing Variant** |  |  |  |  |  |  |  |  |
| No | 677 | 151 | 86 | 0.07 (0.05, 0.09) | 0.18 (0.15, 0.21) | 0.24 (0.2, 0.27) | 1.0 (ref) |  |
| Yes | 46 | 3 | 5 | 0 (0, 0) | 0.05 (0, 0.11) | 0.08 (0, 0.18) | 0.27 (0.09, 0.84) | 0.02 |
| **Cystic Variant** |  |  |  |  |  |  |  |  |
| No | 642 | 148 | 81 | 0.07 (0.05, 0.09) | 0.19 (0.16, 0.22) | 0.25 (0.21, 0.29) | 1.0 (ref) |  |
| Yes | 81 | 6 | 10 | 0.01 (0, 0.04) | 0.04 (0, 0.09) | 0.06 (0, 0.12) | 0.29 (0.13, 0.65) | 0.003 |
| **Vascular Invasion** |  |  |  |  |  |  |  |  |
| No | 558 | 86 | 69 | 0.03 (0.02, 0.05) | 0.1 (0.08, 0.13) | 0.16 (0.12, 0.19) | 1.0 (ref) |  |
| Yes | 151 | 65 | 20 | 0.18 (0.12, 0.25) | 0.45 (0.36, 0.54) | 0.53 (0.43, 0.62) | 4.41 (3.18, 6.12) | < 0.001 |
| **Perineural Invasion** |  |  |  |  |  |  |  |  |
| No | 500 | 73 | 61 | 0.03 (0.02, 0.05) | 0.11 (0.08, 0.14) | 0.16 (0.12, 0.19) | 1.0 (ref) |  |
| Yes | 211 | 80 | 29 | 0.14 (0.09, 0.19) | 0.33 (0.26, 0.4) | 0.4 (0.33, 0.48) | 3.17 (2.31, 4.35) | < 0.001 |
| **Margins** |  |  |  |  |  |  |  |  |
| Negative | 662 | 137 | 78 | 0.06 (0.04, 0.08) | 0.17 (0.14, 0.2) | 0.22 (0.19, 0.26) | 1.0 (ref) |  |
| Positive | 55 | 16 | 12 | 0.13 (0.04, 0.23) | 0.26 (0.14, 0.38) | 0.31 (0.18, 0.45) | 1.47 (0.85, 2.54) | 0.17 |
| **Margins\*** |  |  |  |  |  |  |  |  |
| Negative | 619 | 137 | 76 | 0.06 (0.04, 0.08) | 0.18 (0.14, 0.21) | 0.23 (0.2, 0.27) | 1.0 (ref) |  |
| Positive | 30 | 15 | 7 | 0.21 (0.06, 0.36) | 0.42 (0.24, 0.61) | 0.51 (0.31, 0.71) | 2.78 (1.58, 4.88) | < 0.001 |

\*Excludes patients who had an enucleation procedure. Abbreviations: CIR = Cumulative Incidence of Relapse; Calculated from the date of surgery to last follow-up, date of relapse or death HR =Hazard Ratio, estimated from univariate competing risks regression models.

As with OS, the cumulative incidence of relapse was associated with refined tumor grade. Patients with G1 PanNETs had 5-year cumulative incidence of relapse of 8% (95% CI: 5,11%), those with G2a PanNETs 25% (95% CI: 19, 32%), those with G2b PanNETs 62% (95% CI: 43, 80%), and those with G3 PanNETs 69% (95% CI: 44, 94%) (Table 4). These results again support adding a 10% threshold in the grading system.

### Year and Age of Surgery

A comparison of the distribution of patients diagnosed with stage I, II, III, or IV disease by year of surgery grouped into quintiles revealed a down shift in stage at diagnosis over time (Figure 1E). Over the study period, patients were more likely to be diagnosed with stage I (Odds Ratio (OR) 1.61, 95% CI: 1.34, 1.93, p <0.001), stage II (OR 1.30, 95% CI: 1.08, 1.55, p =0.005), or stage III disease (OR 1.2, 95% CI: 1.00, 1.45, p=0.054) compared to stage IV disease.

Hypothesizing that a down shift in stage over time would be associated with an earlier age at diagnosis, we next examined trends in age at surgery by year of surgery (Supplemental Figure 2). Surprisingly, and perhaps reflecting regional referral practices, we observed a slight increase in the mean age at diagnosis over time, with mean age increasing 1.08 years every five years (95% CI: 0.51, 1.65, p<0.001).

## Discussion

The incidence of PanNETs is increasing.^1–7^ Fortunately, new therapeutic options beyond surgery are available. As clinical trials are designed to define the impact of novel therapies and as clinicians struggle to determine the best therapy for their individual patients, it is important that prognosticators are identified and that their impact on patient survival is quantified accurately.

In this single-institution series of 904 patients with surgically resected PanNETs and extensive follow-up, we provide support for the proposed separation of G2 PanNETs (Ki-67 of 3-≤ 20%) into two grades (G2a for Ki-67 3-<10%, and G2b for Ki-67 of 10%-≤20%).^40^ Here, the proposed grading system clearly stratified the patients for both cumulative incidence of relapse and OS (Figure 1C). Furthermore, analysis of the non-linear relationship between Ki-67 and OS (Figure 1D) revealed a subtle inflection point near a Ki-67 labeling index of 10%. While the separation of patients is clear, we should emphasize that the relationship between Ki-67 and recurrence and OS is generally fairly linear, and all cut-offs are somewhat arbitrary. Furthermore, the division of G2 tumors into two grades (G2a and G2b) may not currently have immediate therapeutic implications.

We also confirm the major prognosticators and refine the magnitude of risk associated with each. The highest HRs for recurrence and OS were associated with metastases at surgery, T stage, N stage, vascular invasion, perineural invasion, and positive margins. Separately analyzing the influence of margin status in a cohort excluding enucleations, as these are performed on low-risk tumors, demonstrates an even greater impact of margin status on prognosis.

Here we show that the serotonin-positive duct-centric sclerosing variant of PanNET comprises 6% of surgically resected PanNETs, and, as has been reported, that patients with this tumor type have an excellent prognosis with a five-year cumulative incidence of relapse of 5% (95% CI: 0%, 11%) and a 10-year OS of 91% (95% CI: 83%, 100%).^32–34,45–47^ This finding has clinical implications as these tumors often involve and stricture the main pancreatic duct causing ductal dilatation and can therefore be detected on imaging as ductal dilatation abruptly ending at a small enhancing mass lesion on computed tomography.^32–34,45–47^ We confirm that insulinomas and cystic PanNETs are associated with a lower risk of recurrence and improved OS.^7,16–20^ Further, in a finding that will improve risk stratification we confirm that that grade, vascular invasion and positive margins are important prognosticators of recurrence and OS in the critical group of patients with T1N0M0 disease.^41^

Finally, we examined changes in stage at diagnosis and patient age at diagnosis over time (Figure 1E and Supplemental Figure 1). The trend towards a lower stage at diagnosis supports the hypothesis that a growing number of asymptomatic patients are being diagnosed incidentally on imaging performed for another indication.^1,43^ However, we did not observe a trend towards younger age at the time of diagnosis over time. In interpreting these trends, one should note that the patients included in this study, who underwent surgery at a high-volume tertiary care center, may not be representative of the entire population of patients with a PanNET.

Weaknesses of the study include the use of two methods to determine the Ki-67 labeling indexes of tumors, although both methods have been used in previous studies.^22,48,53,54^ Finally, OS was used in our initial analyses as it provided, compared to disease-specific survival, the most complete dataset and the exact date of death was clear, while accuracy of the date of recurrence depends on the frequency of surveillance. Because the follow-up on many of the patients was so long, as demonstrated in the cause-specific survival analyses, caution should be taken in interpreting the risk associated with age and gender in OS, as these could be attributable to gender- and age-based differences in mortality in the general population.

Accurate risk stratification of PanNETs will help predict the clinical behavior of these tumors and increase the precision with which future treatment efficacy is assessed. The prognostic significance of grade (Ki-67 index) will help inform the design and interpretation of clinical trials, while other prognostic factors reported here such as histologic subtype or invasive behavior may inform future basic science inquiry into these increasingly common pancreatic tumors.

## Data Availability

All data produced in the present study are available upon reasonable request to the authors.

## Acknowledgements

This work was supported by the Stringer Foundation and the National Institutes of Health (2T32CA193145, EDY).

---

**Supplemental Table 1:** Overall Survival, Patients with M0 Disease: Estimates of survival after surgery for patients with M0 disease, overall and according to patient subgroups defined at the time of surgery.

|  | N | Follow-Up, PY | Median [95% CI] | 10-year OS | 20-year OS | HR | 95% CI | P |
| --- | --- | --- | --- | --- | --- | --- | --- | --- |
| **Whole Cohort** | 767 | 7235 | 22.5 [19.7, 31.2] | 80 [76, 83] | 54 [48, 61] |  |  |  |
| **Age, years** |  |  |  |  |  |  |  |  |
| < 60 | 428 | 4473 | 27.05 [23.89, 40.23+] | 87 [83, 91] | 65 [58, 73] | 1.0 (ref) | - |  |
| 60+ | 339 | 2762 | 15.63 [13.77, 19.69] | 69 [63, 76] | 36 [27, 49] | 2.36 | [1.75, 3.18] | < 0.0001 |
| **Gender** |  |  |  |  |  |  |  |  |
| Female | 367 | 3603 | 26.51 [23.03, 40.23+] | 80 [75, 85] | 65 [58, 73] | 1.0 (ref) | - |  |
| Male | 400 | 3631 | 17.89 [15.63, 23.89] | 80 [75, 84] | 43 [34, 54] | 1.44 | [1.07, 1.93] | 0.02 |
| **Tumor Size** |  |  |  |  |  |  |  |  |
| < 2cm | 322 | 3193 | 27.05 [23.03, 40.23+] | 88 [84, 93] | 67 [57, 78] | 1.0 (ref) | - |  |
| 2cm + | 445 | 4042 | 18.07 [15.1, 23.89] | 73 [68, 78] | 46 [39, 54] | 2.28 | [1.64, 3.18] | < 0.0001 |
| **T Stage** |  |  |  |  |  |  |  |  |
| T1 | 322 | 3193 | 27.05 [23.03, 40.23+] | 88 [84, 93] | 67 [57, 78] | 1.0 (ref) | - |  |
| T2 | 280 | 2557 | 21.16 [17.24, 40.23+] | 77 [71, 83] | 52 [43, 63] | 1.95 | [1.35, 2.81] | 0.0003 |
| T3 | 152 | 1399 | 15.06 [12.16, 40.23+] | 69 [61, 78] | 39 [28, 53] | 2.73 | [1.85, 4.03] | < 0.0001 |
| T4 | 13 | 85 | 9.88 [6.9, 40.23+] | 45 [21, 98] | 0 [21, 98] | 5.4 | [2.3, 12.68] | 0.0001 |
| **N Stage** |  |  |  |  |  |  |  |  |
| N0 | 507 | 4813 | 23.03 [21.16, 40.23+] | 83 [79, 87] | 59 [51, 69] | 1.0 (ref) | - |  |
| Nx or Unknown | 75 | 860 | 31.23 [26.51, 40.23+] | 93 [86, 100] | 81 [68, 95] | 0.48 | [0.24, 0.95] | 0.03 |
| N1 | 185 | 1561 | 12.97 [11.81, 15.98] | 63 [56, 72] | 31 [22, 43] | 2.6 | [1.93, 3.5] | < 0.0001 |
| **Ki-67** |  |  |  |  |  |  |  |  |
| < 2 | 261 | 2811 | 23.03 [19.69, 32.15+] | 84 [79, 89] | 58 [49, 69] | 1.0 (ref) | - |  |
| 2+ | 432 | 3393 | 17.24 [15.45, 32.15+] | 76 [71, 81] | 45 [35, 57] | 1.55 | [1.12, 2.14] | 0.007 |
| **Grade** |  |  |  |  |  |  |  |  |
| G1 | 423 | 4202 | 23.03 [19.69, 32.15+] | 84 [80, 88] | 58 [50, 67] | 1.0 (ref) | - |  |
| G2 | 254 | 1917 | 17.02 [12.97, 23.89] | 72 [65, 80] | 40 [28, 55] | 1.8 | [1.31, 2.48] | 0.0003 |
| G3 | 16 | 84 | 7.24 [2.84, 32.15+] | 32 [12, 82] | 32 [12, 82] | 7.2 | [3.57, 14.53] | < 0.0001 |
| **Grade, Proposed** |  |  |  |  |  |  |  |  |
| G1 | 423 | 4202 | 23.03 [19.69, 32.15+] | 84 [80, 88] | 58 [50, 67] | 1.0 (ref) | - |  |
| G2a | 221 | 1720 | 17.24 [14.42, 23.9] | 75 [67, 83] | 43 [31, 60] | 1.62 | [1.15, 2.27] | 0.006 |
| G2b | 33 | 197 | 10.28 [7.88, 32.15+] | 56 [37, 85] | 14 [2, 80] | 4.02 | [2.17, 7.46] | < 0.0001 |
| G3 | 16 | 84 | 7.24 [2.84, 32.15+] | 32 [12, 82] | 32 [12, 82] | 7.41 | [3.66, 14.97] | < 0.0001 |
| **Insulinoma** |  |  |  |  |  |  |  |  |
| No | 677 | 6301 | 21.16 [17.89, 40.23+] | 78 [74, 82] | 50 [44, 58] | 1.0 (ref) | - |  |
| Yes | 90 | 933 | 27.05 [23.03, 40.23+] | 94 [88, 100] | 82 [71, 96] | 0.36 | [0.19, 0.68] | 0.002 |
| **Sclerosing Variant** |  |  |  |  |  |  |  |  |
| No | 718 | 6786 | 21.52 [18.96, 27.05] | 79 [75, 83] | 53 [47, 60] | 1.0 (ref) | - |  |
| Yes | 49 | 449 | 31.23 [14.56, 40.23+] | 91 [83, 100] | 71 [50, 100] | 0.62 | [0.29, 1.33] | 0.22 |
| **Cystic Variant** |  |  |  |  |  |  |  |  |
| No | 681 | 6401 | 21.4 [18.07, 27.05] | 79 [75, 82] | 52 [46, 60] | 1.0 (ref) | - |  |
| Yes | 86 | 833 | NR | 87 [79, 95] | 68 [52, 89] | 0.67 | [0.39, 1.13] | 0.13 |
| **Vascular Invasion** |  |  |  |  |  |  |  |  |
| No | 594 | 5900 | 23.9 [22.46, 40.23+] | 83 [79, 86] | 60 [53, 67] | 1.0 (ref) | - |  |
| Yes | 157 | 1062 | 14.25 [10.91, 17.24] | 63 [53, 74] | 22 [11, 43] | 2.73 | [1.98, 3.76] | < 0.0001 |
| **Perineural Invasion** |  |  |  |  |  |  |  |  |
| No | 534 | 5242 | 23.9 [22.46, 40.23+] | 83 [79, 87] | 61 [54, 69] | 1.0 (ref) | - |  |
| Yes | 221 | 1787 | 15.1 [12.18, 18.22] | 70 [63, 78] | 31 [21, 48] | 2.32 | [1.73, 3.12] | < 0.0001 |
| **Margins** |  |  |  |  |  |  |  |  |
| Negative | 700 | 6567 | 23.03 [21.16, 40.23+] | 81 [77, 84] | 57 [51, 64] | 1.0 (ref) | - |  |
| Positive | 61 | 577 | 12.55 [11.02, 40.23+] | 64 [51, 80] | 32 [19, 53] | 2.01 | [1.34, 3] | 0.0007 |
| **Margins\*** |  |  |  |  |  |  |  |  |
| Negative | 657 | 6192 | 22.46 [19.66, 40.23+] | 80 [77, 84] | 54 [48, 62] | 1.0 (ref) | - |  |
| Positive | 36 | 310 | 9.88 [8.26, 16.65] | 49 [34, 72] | 16 [6, 43] | 3.03 | [1.95, 4.71] | < 0.0001 |

\*Excludes patients who had an enucleation procedure. Abbreviations: Median survival is in years, 10- and 20-year OS are the respective survival probabilities from the Kaplan-Meier estimate for each subgroup, HR=Hazard Ratio, estimated from univariate Cox proportional hazards regression models, CSS=Cause-specific survival, Calculated from the date of surgery to last follow-up-or death, NR=Median survival not reached Median OS is shown in years, OS=Overall survival; Calculated from the date of surgery to last follow-up or death, PY=Person-years.

**Supplemental Table 2:** Overall Survival, Patients with M0 and N0 Disease: Estimates of survival after surgery for patients with M0 and N0 disease, overall and according to patient subgroups defined at the time of surgery.

|  | N | Follow-Up, PY | Median [95% CI] | 10-year OS | 20-year OS | HR | 95% CI | P |
| --- | --- | --- | --- | --- | --- | --- | --- | --- |
| **Whole Cohort** | 507 | 4814 | 23 [21.2, 36.78+] | 83 [79, 87] | 59 [51, 69] |  |  |  |
| **Age, years** |  |  |  |  |  |  |  |  |
| < 60 | 286 | 2949 | NR | 90 [86, 95] | 70 [60, 82] | 1.0 (ref) | - |  |
| 60+ | 221 | 1864 | 16.81 [14.51, 22.46] | 74 [67, 82] | 42 [29, 60] | 2.78 | [1.83, 4.22] | < 0.0001 |
| **Gender** |  |  |  |  |  |  |  |  |
| Female | 249 | 2387 | NR | 83 [77, 89] | 67 [57, 79] | 1.0 (ref) | - |  |
| Male | 258 | 2426 | 21.52 [17.89, 36.78+] | 84 [79, 90] | 51 [39, 66] | 1.24 | [0.83, 1.86] | 0.3 |
| **Tumor Size** |  |  |  |  |  |  |  |  |
| < 2cm | 251 | 2443 | 23.03 [22.46, 36.78+] | 88 [83, 93] | 65 [52, 80] | 1.0 (ref) | - |  |
| 2cm + | 256 | 2370 | 21.52 [18.07, 36.78+] | 79 [73, 85] | 54 [44, 67] | 1.58 | [1.05, 2.39] | 0.03 |
| **T Stage** |  |  |  |  |  |  |  |  |
| T1 | 251 | 2443 | 23.03 [22.46, 36.78+] | 88 [83, 93] | 65 [52, 80] | 1.0 (ref) | - |  |
| T2 | 170 | 1542 | 21.52 [18.07, 36.78+] | 82 [75, 90] | 55 [41, 74] | 1.38 | [0.86, 2.21] | 0.18 |
| T3 | 81 | 794 | NR | 74 [64, 86] | 54 [40, 72] | 1.88 | [1.11, 3.16] | 0.02 |
| T4 | 5 | 33 | 14.75 [6.9, 36.78+] | 67 [30, 100] | 0 [30, 100] | 4.53 | [1.09, 18.85] | 0.04 |
| **Ki-67** |  |  |  |  |  |  |  |  |
| < 2 | 185 | 1981 | 23.03 [21.52, 32+] | 86 [81, 92] | 62 [51, 77] | 1.0 (ref) | - |  |
| 2+ | 273 | 2152 | 18.07 [14.75, 32+] | 81 [75, 87] | 45 [30, 67] | 1.51 | [0.97, 2.35] | 0.07 |
| **Grade** |  |  |  |  |  |  |  |  |
| G1 | 305 | 2977 | 23.03 [21.52, 32+] | 86 [81, 91] | 63 [52, 75] | 1.0 (ref) | - |  |
| G2 | 146 | 1106 | 16.81 [14.25, 32+] | 79 [70, 89] | 33 [17, 65] | 1.74 | [1.1, 2.75] | 0.02 |
| G3 | 7 | 49 | 8.68 [7.24, 32+] | 34 [8, 100] | 34 [8, 100] | 5.28 | [1.61, 17.3] | 0.006 |
| **Grade, Proposed** |  |  |  |  |  |  |  |  |
| G1 | 305 | 2977 | 23.03 [21.52, 32+] | 86 [81, 91] | 63 [52, 75] | 1.0 (ref) | - |  |
| G2a | 129 | 1004 | 17.02 [14.42, 32+] | 84 [75, 93] | 38 [19, 73] | 1.37 | [0.83, 2.29] | 0.22 |
| G2b | 17 | 102 | 8.54 [5.93, 32+] | 47 [25, 90] | 16 [3, 88] | 6.42 | [2.99, 13.78] | < 0.0001 |
| G3 | 7 | 49 | 8.68 [7.24, 32+] | 34 [8, 100] | 34 [8, 100] | 5.48 | [1.67, 18] | 0.005 |
| **Insulinoma** |  |  |  |  |  |  |  |  |
| No | 456 | 4269 | 22.46 [19.66, 36.78+] | 82 [78, 87] | 55 [46, 66] | 1.0 (ref) | - |  |
| Yes | 51 | 543 | 27.05 [23.03, 36.78+] | 92 [83, 100] | 88 [76, 100] | 0.39 | [0.16, 0.97] | 0.04 |
| **Sclerosing Variant** |  |  |  |  |  |  |  |  |
| No | 467 | 4462 | 23.03 [21.16, 36.78+] | 83 [79, 87] | 58 [50, 68] | 1.0 (ref) | - |  |
| Yes | 40 | 351 | NR | 91 [83, 100] | 81 [63, 100] | 0.61 | [0.22, 1.67] | 0.34 |
| **Cystic Variant** |  |  |  |  |  |  |  |  |
| No | 436 | 4102 | 23.03 [19.66, 36.78+] | 83 [78, 87] | 58 [49, 68] | 1.0 (ref) | - |  |
| Yes | 71 | 711 | NR | 87 [78, 97] | 68 [50, 92] | 0.78 | [0.43, 1.43] | 0.42 |
| **Vascular Invasion** |  |  |  |  |  |  |  |  |
| No | 428 | 4154 | 23.03 [21.52, 32+] | 84 [80, 89] | 60 [51, 71] | 1.0 (ref) | - |  |
| Yes | 68 | 452 | 14.75 [14.25, 32+] | 69 [54, 88] | 46 [25, 85] | 1.92 | [1.08, 3.4] | 0.03 |
| **Perineural Invasion** |  |  |  |  |  |  |  |  |
| No | 390 | 3699 | 23.9 [21.52, 32+] | 84 [79, 89] | 60 [51, 71] | 1.0 (ref) | - |  |
| Yes | 108 | 964 | 21.16 [17.02, 32+] | 81 [72, 90] | 56 [41, 78] | 1.45 | [0.9, 2.31] | 0.12 |
| **Margins** |  |  |  |  |  |  |  |  |
| Negative | 483 | 4625 | 23.9 [21.16, 36.78+] | 85 [81, 89] | 61 [52, 70] | 1.0 (ref) | - |  |
| Positive | 22 | 151 | 9.22 [7.49, 36.78+] | 50 [28, 87] | 25 [8, 77] | 3.94 | [1.97, 7.89] | 0.0001 |
| **Margins\*** |  |  |  |  |  |  |  |  |
| Negative | 469 | 4510 | 23.03 [21.16, 36.78+] | 84 [80, 88] | 60 [51, 70] | 1.0 (ref) | - |  |
| Positive | 16 | 115 | 9.22 [6.9, 36.78+] | 46 [23, 90] | 15 [3, 87] | 4.43 | [2.13, 9.2] | < 0.0001 |

\*Excludes patients who had an enucleation procedure. Abbreviations: Median survival is in years, 10- and 20-year OS are the respective survival probabilities from the Kaplan-Meier estimate for each subgroup, HR=Hazard Ratio, estimated from univariate Cox proportional hazards regression models, CSS=Cause-specific survival, Calculated from the date of surgery to last follow-up-or death, NR=Median survival not reached Median OS is shown in years, OS=Overall survival; Calculated from the date of surgery to last follow-up or death, PY=Person-years.

**Supplemental Table 3:**
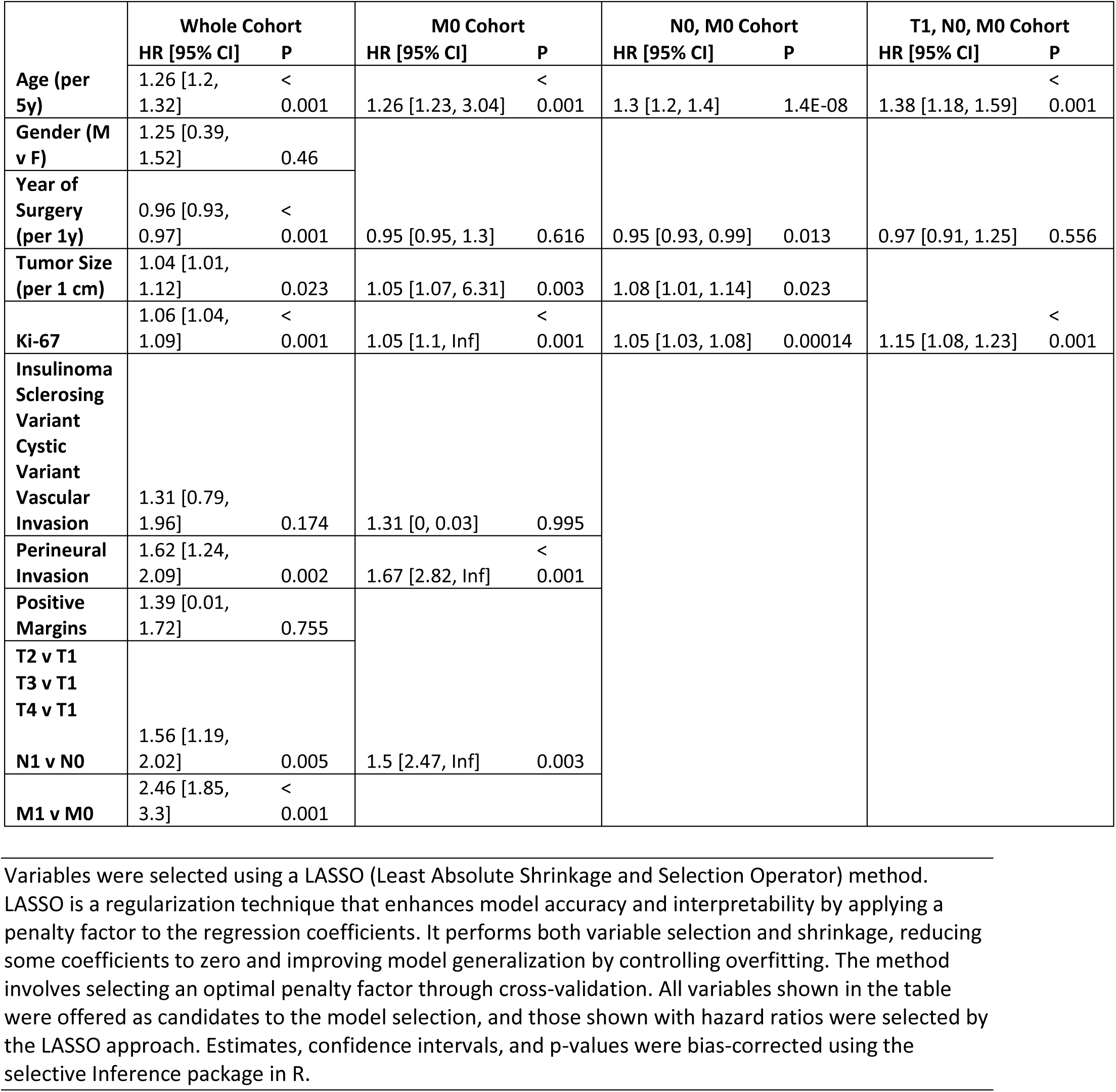
Multivariable Models for Overall Survival: Estimated hazard ratios from multivariable models for overall survival, among patients alive at 30 days after surgery.

**Supplemental Table 4:** Cause-Specific Survival, Patients with M0 Disease: Estimates of survival after surgery for patients with M0 disease, overall and according to patient subgroups defined at the time of surgery.

|  | N | Follow-Up, PY | Median [95% CI] | 10-year CSS | 20-year CSS | HR | 95% CI | P |
| --- | --- | --- | --- | --- | --- | --- | --- | --- |
| **Whole Cohort** | 767 | 7235 | NR | 91 [89, 94] | 80 [75, 86] |  |  |  |
| **Age, years** |  |  |  |  |  |  |  |  |
| < 60 | 428 | 4473 | NR | 93 [90, 96] | 79 [73, 86] | 1.0 (ref) | - |  |
| 60+ | 339 | 2762 | NR | 89 [85, 93] | 84 [78, 91] | 1.29 | [0.79, 2.11] | 0.31 |
| **Gender** |  |  |  |  |  |  |  |  |
| Female | 367 | 3603 | NR | 91 [87, 94] | 85 [80, 91] | 1.0 (ref) | - |  |
| Male | 400 | 3631 | NR | 92 [88, 95] | 73 [64, 84] | 1.3 | [0.8, 2.1] | 0.3 |
| **Tumor Size** |  |  |  |  |  |  |  |  |
| < 2cm | 322 | 3193 | NR | 98 [96, 100] | 96 [93, 100] | 1.0 (ref) | - |  |
| 2cm + | 445 | 4042 | NR | 86 [82, 90] | 69 [62, 78] | 9.84 | [3.96, 24.48] | < 0.0001 |
| **T Stage** |  |  |  |  |  |  |  |  |
| T1 | 322 | 3193 | NR | 98 [96, 100] | 96 [93, 100] | 1.0 (ref) | - |  |
| T2 | 280 | 2557 | NR | 89 [85, 94] | 77 [68, 86] | 7.37 | [2.86, 19.01] | < 0.0001 |
| T3 | 152 | 1399 | 23.89 [18.96, 40.23+] | 80 [72, 88] | 59 [46, 75] | 14.09 | [5.47, 36.32] | < 0.0001 |
| T4 | 13 | 85 | 14.75 [NA, 40.23+] | 80 [52, 100] | 0 [52, 100] | 17.62 | [3.41, 91.09] | 0.0006 |
| **N Stage** |  |  |  |  |  |  |  |  |
| N0 | 507 | 4813 | NR | 95 [93, 98] | 90 [85, 95] | 1.0 (ref) | - |  |
| Nx or Unknown | 75 | 860 | NR | 100 [100, 100] | 100 [100, 100] | 0 | [0, Inf] | 0.99 |
| N1 | 185 | 1561 | 18.96 [15.1, 40.23+] | 75 [67, 84] | 49 [37, 65] | 5.59 | [3.39, 9.22] | < 0.0001 |
| **Ki-67** |  |  |  |  |  |  |  |  |
| < 2 | 261 | 2811 | NR | 97 [94, 100] | 93 [88, 98] | 1.0 (ref) | - |  |
| 2+ | 432 | 3393 | 23.89 [21.16, 32.15+] | 86 [81, 90] | 64 [53, 77] | 5.42 | [2.66, 11.04] | < 0.0001 |
| **Grade** |  |  |  |  |  |  |  |  |
| G1 | 423 | 4202 | NR | 95 [92, 97] | 88 [82, 94] | 1.0 (ref) | - |  |
| G2 | 254 | 1917 | 23.89 [18.96, 32.15+] | 84 [78, 91] | 61 [48, 78] | 3.78 | [2.22, 6.44] | < 0.0001 |
| G3 | 16 | 84 | 8.68 [4.05, 32.15+] | 47 [20, 100] | 47 [20, 100] | 14.11 | [5.22, 38.16] | < 0.0001 |
| **Grade, Proposed** |  |  |  |  |  |  |  |  |
| G1 | 423 | 4202 | NR | 95 [92, 97] | 88 [82, 94] | 1.0 (ref) | - |  |
| G2a | 221 | 1720 | 23.89 [21.16, 32.15+] | 88 [82, 94] | 67 [53, 85] | 3.03 | [1.71, 5.35] | 0.0001 |
| G2b | 33 | 197 | 10.28 [8.54, 32.15+] | 61 [41, 91] | 15 [3, 87] | 12.39 | [5.7, 26.96] | < 0.0001 |
| G3 | 16 | 84 | 8.68 [4.05, 32.15+] | 47 [20, 100] | 47 [20, 100] | 15.12 | [5.55, 41.16] | < 0.0001 |
| **Insulinoma** |  |  |  |  |  |  |  |  |
| No | 677 | 6301 | NR | 90 [88, 93] | 77 [71, 84] | 1.0 (ref) | - |  |
| Yes | 90 | 933 | NR | 97 [93, 100] | 97 [93, 100] | 0.1 | [0.01, 0.76] | 0.03 |
| **Sclerosing Variant** |  |  |  |  |  |  |  |  |
| No | 718 | 6786 | NR | 91 [88, 93] | 80 [74, 85] | 1.0 (ref) | - |  |
| Yes | 49 | 449 | NR | 100 [100, 100] | 86 [63, 100] | 0.24 | [0.03, 1.71] | 0.15 |
| **Cystic Variant** |  |  |  |  |  |  |  |  |
| No | 681 | 6401 | NR | 90 [88, 93] | 78 [72, 84] | 1.0 (ref) | - |  |
| Yes | 86 | 833 | NR | 97 [92, 100] | 97 [92, 100] | 0.23 | [0.06, 0.94] | 0.04 |
| **Vascular Invasion** |  |  |  |  |  |  |  |  |
| No | 594 | 5900 | NR | 94 [92, 96] | 87 [82, 92] | 1.0 (ref) | - |  |
| Yes | 157 | 1062 | 17.24 [14.25, 40.23+] | 75 [65, 85] | 38 [22, 66] | 5.33 | [3.28, 8.65] | < 0.0001 |
| **Perineural Invasion** |  |  |  |  |  |  |  |  |
| No | 534 | 5242 | NR | 96 [93, 98] | 89 [85, 94] | 1.0 (ref) | - |  |
| Yes | 221 | 1787 | 21.16 [17.24, 40.23+] | 80 [74, 87] | 52 [38, 71] | 4.97 | [3.01, 8.2] | < 0.0001 |
| **Margins** |  |  |  |  |  |  |  |  |
| Negative | 700 | 6567 | NR | 92 [89, 94] | 81 [76, 87] | 1.0 (ref) | - |  |
| Positive | 61 | 577 | 23.89 [15.48, 40.23+] | 84 [73, 97] | 64 [47, 88] | 2.26 | [1.18, 4.31] | 0.01 |
| **Margins\*** |  |  |  |  |  |  |  |  |
| Negative | 657 | 6192 | NR | 91 [89, 94] | 80 [74, 86] | 1.0 (ref) | - |  |
| Positive | 36 | 310 | 12.55 [12.03, 40.23+] | 71 [53, 95] | 35 [17, 75] | 4.42 | [2.31, 8.46] | < 0.0001 |

\*Excludes patients who had an enucleation procedure. Patients who died after relapse were considered to have died of their disease. All other deaths were censored.
Abbreviations: Median survival is in years, 10-and 20-year OS are the respective survival probabilities from the Kaplan-Meier estimate for each subgroup, HR=Hazard Ratio, estimated from univariate Cox proportional hazards regression models, CSS=Cause-specific survival, Calculated from the date of surgery to last follow-up-or death, NR=Median survival not reached Median OS is shown in years, OS=Overall survival; Calculated from the date of surgery to last follow-up or death, PY=Person-years.

**Supplemental Table 5:** Cause-Specific Survival, Patients with M0 and N0 Disease: Estimates of survival after surgery for patients with M0 and N0 disease, overall and according to patient subgroups defined at the time of surgery.

|  | N | Follow-Up, PY | Median [95% CI] | 10-year CSS | 20-year CSS | HR | 95% CI | P |
| --- | --- | --- | --- | --- | --- | --- | --- | --- |
| **Whole Cohort** | 507 | 4814 | NR | 95 [93, 98] | 90 [85, 95] |  |  |  |
| **Age, years** |  |  |  |  |  |  |  |  |
| < 60 | 286 | 2949 | NR | 95 [92, 98] | 90 [84, 96] | 1.0 (ref) | - |  |
| 60+ | 221 | 1864 | 23.9 [21.16, 36.78+] | 96 [92, 99] | 89 [81, 99] | 1.3 | [0.57, 2.95] | 0.53 |
| **Gender** |  |  |  |  |  |  |  |  |
| Female | 249 | 2387 | NR | 96 [92, 99] | 91 [84, 97] | 1.0 (ref) | - |  |
| Male | 258 | 2426 | NR | 95 [92, 99] | 89 [82, 96] | 1.08 | [0.48, 2.42] | 0.86 |
| **Tumor Size** |  |  |  |  |  |  |  |  |
| < 2cm | 251 | 2443 | NR | 99 [97, 100] | 96 [92, 100] | 1.0 (ref) | - |  |
| 2cm + | 256 | 2370 | NR | 91 [87, 96] | 83 [75, 91] | 7.18 | [2.14, 24.12] | 0.001 |
| **T Stage** |  |  |  |  |  |  |  |  |
| T1 | 251 | 2443 | NR | 99 [97, 100] | 96 [92, 100] | 1.0 (ref) | - |  |
| T2 | 170 | 1542 | NR | 95 [90, 99] | 93 [87, 99] | 4.12 | [1.09, 15.56] | 0.04 |
| T3 | 81 | 794 | NR | 85 [76, 96] | 70 [55, 88] | 12.65 | [3.56, 44.88] | < 0.0001 |
| T4 | 5 | 33 | 14.75 [NA, 36.78+] | 100 [100, 100] | 0 [100, 100] | 23.12 | [2.39, 223.36] | 0.007 |
| **Ki-67** |  |  |  |  |  |  |  |  |
| < 2 | 185 | 1981 | NR | 98 [96, 100] | 97 [94, 100] | 1.0 (ref) | - |  |
| 2+ | 273 | 2152 | 23.9 [21.16, 32+] | 92 [87, 96] | 74 [60, 90] | 7.85 | [2.31, 26.7] | 0.001 |
| **Grade** |  |  |  |  |  |  |  |  |
| G1 | 305 | 2977 | NR | 96 [94, 99] | 93 [89, 98] | 1.0 (ref) | - |  |
| G2 | 146 | 1106 | 21.16 [21.16, 32+] | 92 [86, 99] | 70 [53, 92] | 3.95 | [1.72, 9.07] | 0.001 |
| G3 | 7 | 49 | 8.68 [8.68, 32+] | 50 [13, 100] | 50 [13, 100] | 7.86 | [0.97, 63.73] | 0.05 |
| **Grade, Proposed** |  |  |  |  |  |  |  |  |
| G1 | 305 | 2977 | NR | 96 [94, 99] | 93 [89, 98] | 1.0 (ref) | - |  |
| G2a | 129 | 1004 | 23.9 [21.16, 32+] | 97 [94, 100] | 79 [61, 100] | 2.36 | [0.89, 6.23] | 0.08 |
| G2b | 17 | 102 | 11.35 [7.88, 32+] | 56 [31, 100] | 19 [3, 100] | 25.42 | [8.67, 74.58] | < 0.0001 |
| G3 | 7 | 49 | 8.68 [8.68, 32+] | 50 [13, 100] | 50 [13, 100] | 8.88 | [1.08, 73.04] | 0.04 |
| **Insulinoma** |  |  |  |  |  |  |  |  |
| No | 456 | 4269 | NR | 95 [93, 98] | 89 [84, 94] | 1.0 (ref) | - |  |
| Yes | 51 | 543 | NR | 95 [87, 100] | 95 [87, 100] | 0.36 | [0.05, 2.71] | 0.32 |
| **Sclerosing Variant** |  |  |  |  |  |  |  |  |
| No | 467 | 4462 | NR | 95 [92, 98] | 89 [84, 94] | 1.0 (ref) | - |  |
| Yes | 40 | 351 | NR | 100 [100, 100] | 100 [100, 100] | 0 | [0, Inf] | > 0.99 |
| **Cystic Variant** |  |  |  |  |  |  |  |  |
| No | 436 | 4102 | NR | 95 [92, 98] | 88 [82, 94] | 1.0 (ref) | - |  |
| Yes | 71 | 711 | NR | 98 [94, 100] | 98 [94, 100] | 0.23 | [0.03, 1.73] | 0.15 |
| **Vascular Invasion** |  |  |  |  |  |  |  |  |
| No | 428 | 4154 | NR | 96 [94, 99] | 92 [88, 97] | 1.0 (ref) | - |  |
| Yes | 68 | 452 | 21.16 [14.25, 32+] | 83 [70, 99] | 56 [31, 99] | 5.19 | [2.21, 12.19] | 0.0002 |
| **Perineural Invasion** |  |  |  |  |  |  |  |  |
| No | 390 | 3699 | NR | 97 [95, 99] | 91 [85, 97] | 1.0 (ref) | - |  |
| Yes | 108 | 964 | NR | 91 [84, 98] | 85 [75, 96] | 2.46 | [1.05, 5.77] | 0.04 |
| **Margins** |  |  |  |  |  |  |  |  |
| Negative | 483 | 4625 | NR | 96 [94, 98] | 90 [86, 95] | 1.0 (ref) | - |  |
| Positive | 22 | 151 | NR | 82 [62, 100] | 55 [23, 100] | 5.04 | [1.48, 17.19] | 0.01 |
| **Margins\*** |  |  |  |  |  |  |  |  |
| Negative | 469 | 4510 | NR | 96 [93, 98] | 90 [85, 95] | 1.0 (ref) | - |  |
| Positive | 16 | 115 | 12.08 [12.08, 36.78+] | 77 [52, 100] | 38 [9, 100] | 6.42 | [1.88, 21.88] | 0.003 |

**Table 6:** Cumulative Incidence of Relapse: Estimates of cumulative probability of relapse at 2, 5 and 10 years after surgery, accounting for death before relapse as a competing event, for the N0M0 cohort and according to patient subgroups defined at the time of surgery.

|  | N | N Relapses | N Deaths | CIR, 2y | CIR, 5y | CIR, 10y | HR (95% CI) | P |
| --- | --- | --- | --- | --- | --- | --- | --- | --- |
| **Whole Cohort** | 507 | 71 | 57 | 0.03 (0.01, 0.05) | 0.11 (0.08, 0.14) | 0.16 (0.13, 0.2) |  |  |
| **Age, years** |  |  |  |  |  |  |  |  |
| < 60 | 276 | 42 | 22 | 0.03 (0.01, 0.06) | 0.12 (0.08, 0.16) | 0.17 (0.12, 0.22) | 1.0 (ref) |  |
| 60+ | 205 | 29 | 35 | 0.02 (0, 0.05) | 0.1 (0.06, 0.15) | 0.15 (0.1, 0.21) | 0.94 (0.59, 1.51) | 0.81 |
| **Gender** |  |  |  |  |  |  |  |  |
| Female | 239 | 35 | 25 | 0.03 (0.01, 0.06) | 0.11 (0.06, 0.15) | 0.16 (0.1, 0.21) | 1.0 (ref) |  |
| Male | 242 | 36 | 32 | 0.03 (0.01, 0.05) | 0.12 (0.08, 0.16) | 0.17 (0.12, 0.22) | 1 (0.63, 1.58) | 0.99 |
| **Tumor Size** |  |  |  |  |  |  |  |  |
| < 2cm | 241 | 15 | 29 | 0.01 (0, 0.02) | 0.05 (0.02, 0.08) | 0.06 (0.03, 0.1) | 1.0 (ref) |  |
| 2cm + | 240 | 56 | 28 | 0.05 (0.02, 0.08) | 0.18 (0.13, 0.24) | 0.27 (0.2, 0.34) | 4.54 (2.58, 7.98) | < 0.001 |
| **T Stage** |  |  |  |  |  |  |  |  |
| T1 | 241 | 15 | 29 | 0.01 (0, 0.02) | 0.05 (0.02, 0.08) | 0.06 (0.03, 0.1) | 1.0 (ref) |  |
| T2 | 160 | 27 | 19 | 0.04 (0.01, 0.07) | 0.13 (0.07, 0.19) | 0.2 (0.13, 0.28) | 3.25 (1.74, 6.07) | < 0.001 |
| T3 | 75 | 27 | 8 | 0.08 (0.02, 0.14) | 0.27 (0.17, 0.38) | 0.38 (0.26, 0.5) | 7.04 (3.75, 13.22) | < 0.001 |
| T4 | 5 | 2 | 1 | 0 (0, 0) | 0.5 (0, 1.09) |  | 10.14 (2.15, 47.77) | 0.003 |
| **Ki-67** |  |  |  |  |  |  |  |  |
| < 2 | 175 | 11 | 28 | 0.02 (0, 0.04) | 0.04 (0.01, 0.07) | 0.08 (0.03, 0.12) | 1.0 (ref) |  |
| 2+ | 264 | 53 | 23 | 0.03 (0.01, 0.05) | 0.17 (0.12, 0.22) | 0.24 (0.18, 0.3) | 4.19 (2.18, 8.06) | < 0.001 |
| **Grade** |  |  |  |  |  |  |  |  |
| G1 | 290 | 21 | 34 | 0.02 (0, 0.04) | 0.06 (0.03, 0.08) | 0.08 (0.05, 0.12) | 1.0 (ref) |  |
| G2 | 142 | 38 | 15 | 0.04 (0.01, 0.07) | 0.21 (0.13, 0.29) | 0.35 (0.25, 0.46) | 5.05 (2.94, 8.66) | < 0.001 |
| G3 | 7 | 5 | 2 | 0 (0, 0) | 0.71 (0.3, 1.13) |  | 16.35 (5.77, 46.33) | < 0.001 |
| **Grade, Proposed** |  |  |  |  |  |  |  |  |
| G1 | 290 | 21 | 34 | 0.02 (0, 0.04) | 0.06 (0.03, 0.08) | 0.08 (0.05, 0.12) | 1.0 (ref) |  |
| G2a | 125 | 29 | 13 | 0.03 (0, 0.05) | 0.17 (0.1, 0.25) | 0.3 (0.2, 0.41) | 4.24 (2.41, 7.47) | < 0.001 |
| G2b | 17 | 9 | 2 | 0.13 (0, 0.31) | 0.49 (0.21, 0.76) | 0.71 (0.37, 1.05) | 13.41 (5.97, 30.14) | < 0.001 |
| G3 | 7 | 5 | 2 | 0 (0, 0) | 0.71 (0.3, 1.13) |  | 16.44 (5.77, 46.82) | < 0.001 |
| **Insulinoma** |  |  |  |  |  |  |  |  |
| No | 436 | 67 | 54 | 0.03 (0.01, 0.05) | 0.11 (0.08, 0.14) | 0.17 (0.13, 0.21) | 1.0 (ref) |  |
| Yes | 45 | 4 | 3 | 0.02 (0, 0.07) | 0.11 (0.01, 0.22) | 0.11 (0.01, 0.22) | 0.59 (0.22, 1.63) | 0.31 |
| **Sclerosing Variant** |  |  |  |  |  |  |  |  |
| No | 443 | 70 | 53 | 0.03 (0.02, 0.05) | 0.12 (0.09, 0.16) | 0.17 (0.13, 0.21) | 1.0 (ref) |  |
| Yes | 38 | 1 | 4 | 0 (0, 0) | 0 (0, 0) | 0.04 (0, 0.13) | 0.16 (0.02, 1.11) | 0.06 |
| **Cystic Variant** |  |  |  |  |  |  |  |  |
| No | 414 | 68 | 49 | 0.03 (0.02, 0.05) | 0.13 (0.09, 0.16) | 0.18 (0.14, 0.23) | 1.0 (ref) |  |
| Yes | 67 | 3 | 8 | 0 (0, 0) | 0.02 (0, 0.05) | 0.04 (0, 0.09) | 0.25 (0.08, 0.77) | 0.02 |
| **Vascular Invasion** |  |  |  |  |  |  |  |  |
| No | 405 | 45 | 50 | 0.01 (0, 0.02) | 0.08 (0.05, 0.1) | 0.13 (0.09, 0.16) | 1.0 (ref) |  |
| Yes | 66 | 24 | 5 | 0.15 (0.06, 0.24) | 0.37 (0.24, 0.51) | 0.43 (0.28, 0.57) | 5.29 (3.21, 8.72) | < 0.001 |
| **Perineural Invasion** |  |  |  |  |  |  |  |  |
| No | 367 | 43 | 42 | 0.03 (0.01, 0.04) | 0.09 (0.06, 0.13) | 0.13 (0.09, 0.17) | 1.0 (ref) |  |
| Yes | 105 | 27 | 14 | 0.05 (0.01, 0.09) | 0.18 (0.1, 0.26) | 0.28 (0.18, 0.37) | 2.27 (1.41, 3.67) | < 0.001 |
| **Margins** |  |  |  |  |  |  |  |  |
| Negative | 457 | 65 | 51 | 0.03 (0.01, 0.04) | 0.11 (0.08, 0.14) | 0.16 (0.12, 0.19) | 1.0 (ref) |  |
| Positive | 22 | 6 | 6 | 0.09 (0, 0.21) | 0.25 (0.05, 0.45) | 0.32 (0.1, 0.54) | 2.32 (0.96, 5.63) | 0.06 |
| **Margins\*** |  |  |  |  |  |  |  |  |
| Negative | 443 | 65 | 51 | 0.03 (0.01, 0.04) | 0.11 (0.08, 0.14) | 0.16 (0.12, 0.2) | 1.0 (ref) |  |
| Positive | 16 | 5 | 5 | 0.06 (0, 0.18) | 0.28 (0.04, 0.53) | 0.36 (0.09, 0.63) | 2.53 (0.97, 6.6) | 0.06 |

\*Excludes patients who had an enucleation procedure. Abbreviations: CIR = Cumulative Incidence of Relapse; Calculated from the date of surgery to last follow-up, date of relapse or death HR =Hazard Ratio, estimated from univariate competing risks regression models.

**Table 7:** Cumulative Incidence of Relapse: Estimates of cumulative probability of relapse at 2, 5 and 10 years after surgery, accounting for death before relapse as a competing event, for the T1N0M0 cohort and according to patient subgroups defined at the time of surgery.

|  | N | N<br>Relapses | N<br>Deaths | CIR, 2y | CIR, 5y | CIR, 10y | HR (95% CI) | P |
| --- | --- | --- | --- | --- | --- | --- | --- | --- |
| **Whole Cohort** | 251 | 15 | 29 | 0.01 (0, 0.02) | 0.05 (0.02, 0.08) | 0.06 (0.03, 0.1) |  |  |
| **Age, years** |  |  |  |  |  |  |  |  |
| < 60 | 138 | 8 | 14 | 0.01 (0, 0.02) | 0.03 (0, 0.06) | 0.06 (0.02, 0.11) | 1.0 (ref) |  |
| 60+ | 103 | 7 | 15 | 0.01 (0, 0.03) | 0.07 (0.01, 0.12) | 0.07 (0.01, 0.12) | 1.26 (0.46, 3.45) | 0.66 |
| **Gender** |  |  |  |  |  |  |  |  |
| Female | 126 | 10 | 16 | 0.01 (0, 0.02) | 0.06 (0.01, 0.1) | 0.08 (0.03, 0.13) | 1.0 (ref) |  |
| Male | 115 | 5 | 13 | 0.01 (0, 0.03) | 0.04 (0, 0.07) | 0.05 (0.01, 0.09) | 0.53 (0.18, 1.54) | 0.24 |
| **Ki-67** |  |  |  |  |  |  |  |  |
| < 2 | 107 | 4 | 18 | 0.02 (0, 0.04) | 0.04 (0, 0.08) | 0.04 (0, 0.08) | 1.0 (ref) |  |
| 2+ | 109 | 10 | 8 | 0 (0, 0) | 0.07 (0.02, 0.12) | 0.11 (0.04, 0.18) | 2.82 (0.85, 9.34) | 0.09 |
| **Grade** |  |  |  |  |  |  |  |  |
| G1 | 169 | 5 | 21 | 0.01 (0, 0.03) | 0.03 (0, 0.06) | 0.03 (0, 0.06) | 1.0 (ref) |  |
| G2 | 46 | 9 | 4 | 0 (0, 0) | 0.15 (0.03, 0.28) | 0.27 (0.1, 0.44) | 8.75 (2.77, 27.6) | < 0.001 |
| **Grade, Proposed** |  |  |  |  |  |  |  |  |
| G1 | 169 | 5 | 21 | 0.01 (0, 0.03) | 0.03 (0, 0.06) | 0.03 (0, 0.06) | 1.0 (ref) |  |
| G2a | 44 | 8 | 3 | 0 (0, 0) | 0.13 (0.01, 0.26) | 0.26 (0.09, 0.43) | 8.05 (2.49, 26.02) | < 0.001 |
| G2b | 2 | 1 | 1 | 0 (0, 0) |  |  | 28.63 (2.35, 348.9) | 0.009 |
| **Insulinoma** |  |  |  |  |  |  |  |  |
| No | 210 | 13 | 27 | 0.01 (0, 0.02) | 0.04 (0.01, 0.07) | 0.06 (0.03, 0.1) | 1.0 (ref) |  |
| Yes | 31 | 2 | 2 | 0 (0, 0) | 0.08 (0, 0.19) | 0.08 (0, 0.19) | 1.12 (0.25, 4.94) | 0.88 |
| **Sclerosing Variant** |  |  |  |  |  |  |  |  |
| No | 206 | 14 | 25 | 0.01 (0, 0.02) | 0.05 (0.02, 0.09) | 0.07 (0.03, 0.1) | 1.0 (ref) |  |
| Yes | 35 | 1 | 4 | 0 (0, 0) | 0 (0, 0) | 0.05 (0, 0.14) | 0.44 (0.06, 3.32) | 0.43 |
| **Cystic Variant** |  |  |  |  |  |  |  |  |
| No | 209 | 14 | 23 | 0.01 (0, 0.02) | 0.05 (0.02, 0.08) | 0.07 (0.03, 0.11) | 1.0 (ref) |  |
| Yes | 32 | 1 | 6 | 0 (0, 0) | 0.03 (0, 0.09) | 0.03 (0, 0.09) | 0.4 (0.05, 3.13) | 0.38 |
| **Vascular Invasion** |  |  |  |  |  |  |  |  |
| No | 221 | 10 | 25 | 0 (0, 0.01) | 0.03 (0, 0.05) | 0.04 (0.01, 0.07) | 1.0 (ref) |  |
| Yes | 12 | 4 | 3 | 0.08 (0, 0.25) | 0.36 (0.05, 0.67) | 0.36 (0.05, 0.67) | 10.18 (3.19, 32.49) | < 0.001 |
| **Perineural Invasion** |  |  |  |  |  |  |  |  |
| No | 190 | 9 | 22 | 0.01 (0, 0.03) | 0.04 (0.01, 0.07) | 0.05 (0.02, 0.08) | 1.0 (ref) |  |
| Yes | 44 | 6 | 7 | 0 (0, 0) | 0.08 (0, 0.17) | 0.15 (0.02, 0.27) | 2.89 (1.04, 8) | 0.04 |
| **Margins** |  |  |  |  |  |  |  |  |
| Negative | 232 | 14 | 26 | 0 (0, 0.01) | 0.04 (0.02, 0.07) | 0.06 (0.03, 0.1) | 1.0 (ref) |  |
| Positive | 8 | 1 | 3 | 0.12 (0, 0.37) | 0.12 (0, 0.37) | 0.12 (0, 0.37) | 2.53 (0.3, 21.68) | 0.4 |
| **Margins\*** |  |  |  |  |  |  |  |  |
| Negative | 223 | 14 | 26 | 0 (0, 0.01) | 0.05 (0.02, 0.08) | 0.06 (0.03, 0.1) | 1.0 (ref) |  |
| Positive | 4 | 0 | 3 | 0 (0, 0) | 0 (0, 0) |  | 0 (0, 0) | < 0.001 |

\*Excludes patients who had an enucleation procedure. Abbreviations: CIR = Cumulative Incidence of Relapse; Calculated from the date of surgery to last follow-up, date of relapse or death HR =Hazard Ratio, estimated from univariate competing risks regression models.

**Supplemental Figure 1:**
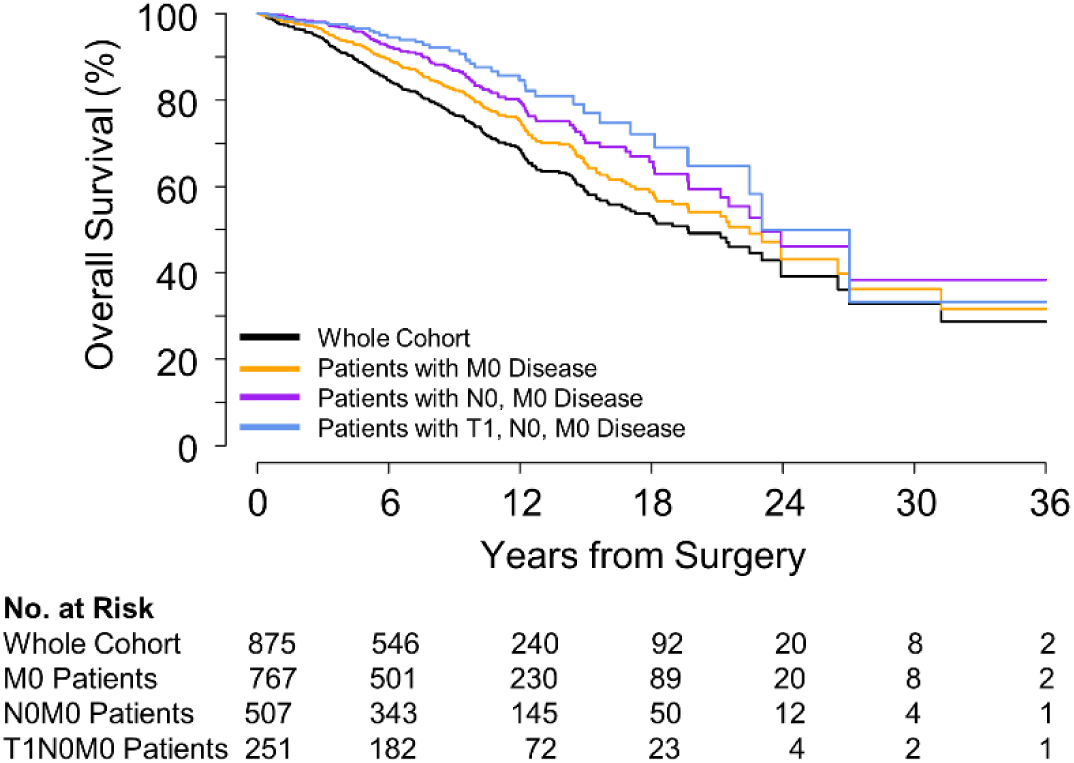
Kaplan Meyer Curve for entire cohort and for the subset of cohort that is M0; N0, M0, and T1, N0, M0.

**Supplemental Figure 2:**
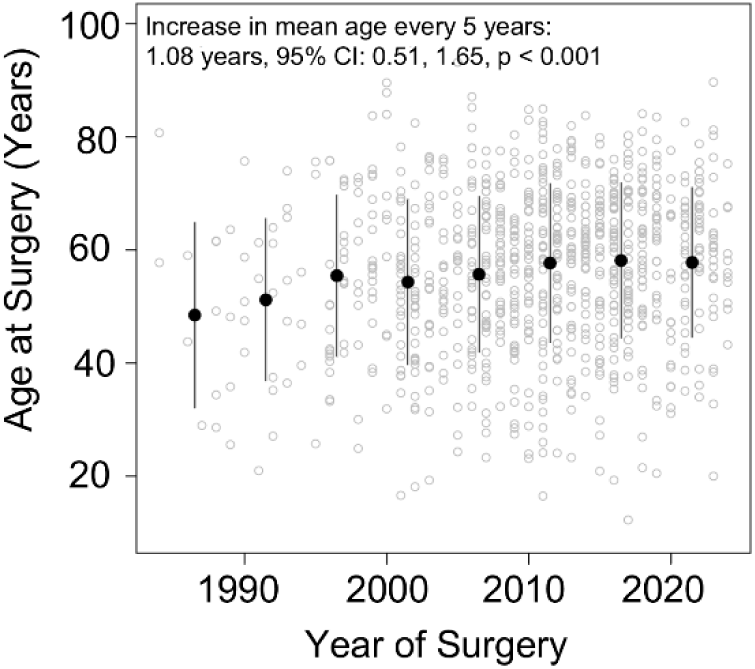
Scatterplot of age at surgery (y-axis) by year of surgery (x-axis). Grey dots represent individual patient data and black points and bars are the mean and standard deviation of age in five-year intervals, starting with 1984 to 1990, 1991 to 1995, and so on. Change in mean age with every five years is estimated from a simple linear regression model.

## Notes

### Competing Interest Statement

The authors have declared no competing interest.

### Funding Statement

This study was funded by Stringer Foundation, and Doug and Julie Ostrover.

### Author Declarations

This study was approved by the Johns Hopkins University Institutional Review Board.

